# Adipose tissue distribution from body MRI is associated with cross-sectional and longitudinal brain age in adults

**DOI:** 10.1101/2021.04.08.21255106

**Authors:** Dani Beck, Ann-Marie G. de Lange, Dag Alnæs, Ivan I. Maximov, Mads L. Pedersen, Olof Dahlqvist Leinhard, Jennifer Linge, Rozalyn Simon, Geneviève Richard, Kristine M. Ulrichsen, Erlend S. Dørum, Knut K. Kolskår, Anne-Marthe Sanders, Adriano Winterton, Tiril P. Gurholt, Tobias Kaufmann, Nils Eiel Steen, Jan Egil Nordvik, Ole A. Andreassen, Lars T. Westlye

**Author notes:** Corresponding authors: Dani Beck & Lars T. Westlye, Department of Psychology, University of Oslo, PoBox 1094 Blindern, 0317 OSLO, Norway.

## Abstract

There is an intimate body-brain connection in ageing, and obesity is a key risk factor for poor cardiometabolic health and neurodegenerative conditions. Although research has demonstrated deleterious effects of obesity on brain structure and function, the majority of studies have used conventional measures such as waist-to-hip ratio, waist circumference, and body mass index. While sensitive to gross features of body composition, such global anthropomorphic features fail to describe regional differences in body fat distribution and composition, and to determine visceral adiposity, which is related to a range of metabolic conditions. In this mixed cross-sectional and longitudinal design (interval mean and standard deviation = 19.7 ± 0.5 months), including 790 healthy individuals (mean (range) age = 46.7 (18-94) years, 53% women), we investigated cross-sectional body magnetic resonance imaging (MRI, n = 286) measures of adipose tissue distribution in relation to longitudinal brain structure using MRI-based morphometry and diffusion tensor imaging (DTI). We estimated tissue-specific brain age at two time points and performed Bayesian multilevel modelling to investigate the associations between adipose measures at follow-up and brain age gap (BAG) at baseline and follow-up. We also tested for interactions between BAG and both time and age on each adipose measure. The results showed credible associations between T1-based BAG and liver fat, muscle fat infiltration (MFI), and weight-to-muscle ratio (WMR), indicating older-appearing brains in people with higher measures of adipose tissue. Longitudinal evidence supported interaction effects between time and MFI and WMR on T1-based BAG, indicating accelerated ageing over the course of the study period in people with higher measures of adipose tissue. The results show that specific measures of fat distribution are associated with brain ageing and that different compartments of adipose tissue may be differentially linked with increased brain ageing, with potential to identify key processes involved in age-related transdiagnostic disease processes.

## 1. Introduction

An increasing body of evidence supports close body-brain connections in ageing, with cardiovascular disease (CVD), cognitive decline, and dementia sharing various cardiometabolic risk factors (Qiu & Fratiglioni, 2015). Among these, obesity subsists as a key risk factor (Bhupathiraju & Hu, 2016; Luppino et al., 2010), with evidence extending the links to include mental disorders (Bahrami et al., 2020; Ditmars et al., 2021; Luppino et al., 2010; Perry et al., 2021; Quintana et al., 2017; Rajan & Menon, 2017; Ringen et al., 2018; Scott et al., 2008) and age-related neurocognitive and neurological conditions including dementia and stroke (Anstey et al., 2011; Strazzullo et al., 2010).

Higher adipose tissue levels as indexed by waist circumference (WC), waist-to-hip ratio (WHR), body mass index (BMI), and increased subcutaneous (ASAT) and visceral (VAT) adipose tissue measures have all been associated with global brain volume decreases (Debette & Markus, 2010; Gunstad et al., 2005; Mulugeta et al., 2021; Ward et al., 2005). Moreover, regional findings have consistently shown negative associations between obesity and brain grey matter volume (Gurholt et al., 2020; Pannacciulli et al., 2006; Taki et al., 2008; Walther et al., 2010) and white matter microstructure, including reduced white matter tract coherence (Friedman et al., 2014; Willette & Kapogiannis, 2015), white matter integrity (Marks et al., 2011; Stanek et al., 2011; Xu et al., 2013), microstructural changes in childhood (Rapuano et al., 2020), and increased axonal and myelin damage (Mueller et al., 2011; Xu et al., 2013) based on diffusion MRI. White matter volumetric studies have revealed less consistent findings, reporting both positive (Walther et al., 2010), negative (Raji et al., 2010) and no (Gunstad et al., 2005) significant associations between brain white matter volume and adiposity.

While there is a wealth of research focusing on conventional anthropomorphic measures such as BMI, not all individuals with a higher BMI have the same disease risks (Mulugeta et al., 2021). A study assessing 27,000 individuals from 52 countries identified abdominal obesity as one of nine key risk factors that accounted for most of the risk of myocardial infarction worldwide (Yusuf et al., 2004). However, while some obese individuals develop health problems such as lipid abnormalities and type 2 diabetes (Lacobini et al., 2019), others are metabolically healthy. Partly motivated by this heterogeneity and complexity of fat distribution and cardiometabolic health, body MRI has recently emerged as a novel opportunity to investigate adipose tissue distribution beyond anthropomorphic measures (Linge et al., 2018, 2019, 2020, 2021).

Research utilising body MRI has found associations between visceral adiposity tissue (VAT) and muscle fat infiltration (MFI) and coronary heart disease (CHD) and type 2 diabetes (T2D) (Linge et al., 2018). Moreover, higher liver fat has been associated with T2D and lower liver fat with CHD (Linge et al., 2018). Cross-sectional analyses investigating body and brain MRI associations have reported negative associations between liver fat, MFI and cerebral cortical thickness while thigh muscle volume was positively associated with brain stem and accumbens volumes (Gurholt et al., 2020).

Advanced multimodal brain MRI provides a wealth of information reflecting structural and functional characteristics of the brain. Brain age prediction using machine learning and a combination of MRI features provides a reliable approach for reducing the complexity and dimensionality of imaging data. The difference between the brain-predicted age and an individual’s chronological age, also referred to as the brain age gap (BAG), can be used to assess deviations from expected age trajectories, with potential utility in studies of brain disorders and ageing (Cole et al., 2017; Kaufmann et al., 2019). This has clear clinical implications for patient groups, where studies have reported larger brain age gaps in patients with various neurological and psychiatric disorders (Han et al., 2020; Høgestøl et al., 2019; Kaufmann et al., 2019; Pardoe et al., 2017; Sone et al., 2019; Tønnesen et al., 2020).

Recent evidence has demonstrated that the rate of brain ageing may be dependent on a range of life events and lifestyle factors (Cole, 2020; Sanders et al., 2021), and characteristics related to cardiovascular health and obesity, including WHR and BMI (Beck et al., 2021b; de Lange et al., 2020; Franke et al., 2014; Kolbeinsson et al., 2020; Kolenic et al., 2018; Ronan et al., 2016).

In the current study, our primary aim was to identify interactions between adipose tissue measures based on body MRI and tissue specific (DTI and T1-weighted) measures of brain age. We investigated cross-sectional associations of tissue specific BAG and detailed adipose tissue measures (body composition) and, for comparison, conventional anthropomorphic measures (BMI and WHR) used in a recent study (Beck et al., 2021b). Next, we tested for associations between longitudinal brain age and body composition at follow-up and investigated associations between each adiposity measure and longitudinal BAG. Adopting a Bayesian statistical framework, we hypothesised that higher abdominal fat ratio, weight-to-muscle ratio, total (abdominal) adipose volume, visceral fat index, muscle fat infiltration, and liver fat percentage would be associated with older appearing brains, with stronger associations in the body MRI measures than for traditional anthropomorphic features. Further, we hypothesised that indices of obesity are associated with accelerated brain ageing as reflected in larger changes in BAG between baseline and follow-up.

## 2. Material and methods

### 2.1. Sample description

Two integrated studies - the Thematically Organised Psychosis (TOP) (Tønnesen et al., 2018) and StrokeMRI (Richard et al., 2018) - formed the initial sample, which included 1130 datasets from 832 healthy participants. All procedures were conducted in line with the Declaration of Helsinki and the study has been approved by the Norwegian Regional Committees for Medical and Health Research Ethics (REC). All participants provided written informed consent, and exclusion criteria included neurological and mental disorders, and previous head trauma.

Following the removal of 68 datasets after quality checking (QC) of the MRI data (see section 2.5), the final sample included 1062 brain MRI datasets collected from 790 healthy participants aged 18-94 years (mean ± standard deviation (SD) at baseline: 46.8 ± 16.3). This included longitudinal data (two time-points with 19.7 months interval, on average (min = 9.8, max = 35.6) from 272 participants. Of the 790 included participants, body MRI data was available from a subgroup of 286 participants, with age range 19-86 (mean = 57.6, SD = 15.6). Demographic information is summarised in Table 1 and Figure 1.

**Table 1.** Descriptive characteristics of the study sample.

|  | **Baseline**<br>**brain MRI** | **Follow-up**<br>**brain MRI** | **Body MRI** | **Males at**<br>**baseline** | **Females at**<br>**baseline** | **Males at**<br>**follow-up** | **Females at**<br>**follow-up** |
| --- | --- | --- | --- | --- | --- | --- | --- |
| *N* subjects | 790 | 272 | 286 | 372 (47.1%) | 418 (52.9%) | 106 (39%) | 166 (61%) |
| **Sex (Males/Females)** | 372/418 | 106/166 | 110/176 | 372/0 | 0/418 | 106/0 | 0/166 |
| **Age (mean ± SD)** | 46.7 ± 16.3 | 56.9 ± 15.0 | 57.6 ± 15.6 | 45.4 ± 16.3 | 48.0 ± 16.3 | 56.7 ± 16.9 | 57.1 ± 13.6 |
| **Predicted Age T1** | 46.6 ± 17.4 | 56.7 ± 16.8 | 57.8 ± 17.7 | 44.6 ± 16.8 | 48.4 ± 17.3 | 55.3 ± 17.9 | 57.6 ± 16.0 |
| **Predicted Age DTI** | 46.5 ± 16.9 | 57.0 ± 15.9 | 58.2 ± 16.9 | 46.9 ± 16.7 | 47.1 ± 17.0 | 57.7 ± 17.3 | 56.6 ± 15.0 |
| **BAG T1** | -0.15 ± 6.4 | -0.23 ± 6.9 | 0.2 ± 6.9 | -0.8 ± 5.9 | 0.5 ± 6.9 | -1.41 ± 6.4 | 0.53 ± 7.2 |
| **BAG DTI** | -0.2 ± 5.1 | 0.05 ± 0.9 | 0.5 ± 5.4 | 0-5 ± 5.1 | -0.9 ± 5.0 | 0.92 ± 5.0 | -0.51 ± 5.4 |
| **BMI, kg/m<sup>2</sup>** | 25.2 ± 4.06 | 25.0 ± 3.7 |  | 25.6 ± 3.7 | 24.8 ± 4.3 | 25.3 ± 2.9 | 25.3 ± 4.4 |
| **Waist-to-hip ratio (WHR)** | 0.87 ± 0.09 | 0.9 ± 0.09 |  | 0.91 ± 0.1 | 0.83 ± 0.1 | 0.93 ± 0.07 | 0.86 ± 0.08 |
| **Visceral adipose tissue (VAT)**<br>**(min-max)** |  |  | 2.7 (0.4-9.2) | 3.7 (0.6-9.2) | 2.2 (0.4-5.5) |  |  |
| **Abdominal subcutaneous**<br>**adipose tissue (ASAT)** |  |  | 6.3 (1.1-19.7) | 4.9 (1.5-13.4) | 7.1 (1.1-19.8) |  |  |
| **Thigh muscle volume (TMV)** |  |  | 2.6 (0.6-1.5) | 3.3 (2.3-4.2) | 2.2 (1.5-3.3) |  |  |
| **Weight-to-muscle ratio (WMR)** |  |  | 29.3 (12.0-82.5) | 25.6 (19.4-33.9) | 31.7 (12.0-82.5) |  |  |
| **Liver PDFF (Liver fat)** |  |  | 4.3 (1.1-29.8) | 4.9 (1.1-29.8) | 3.9 (1.24-27.8) |  |  |
| **Fat ratio** |  |  | 74.5 (34.9-90.5) | 70.0 (34.9-84.7) | 77.5 (42.7-90.5) |  |  |
| **Visceral abdominal adipose**<br>**tissue index (VAT index)** |  |  | 0.9 (0.14-2.9) | 1.1 (0.2-2.9) | 0.8 (0.14-2.0) |  |  |
| **Total abdominal adipose tissue**<br>**index (Total adipose)** |  |  | 3.0 (0.51-8.5) | 2.6 (0.57-5.3) | 3.3 (0.51-8.5) |  |  |
| **Muscle fat infiltration (MFI)** |  |  | 7.6 (3.6-17.2) | 6.9 (3.6-12.8) | 8.3 (4.5-17.2) |  |  |

**Figure 1.**
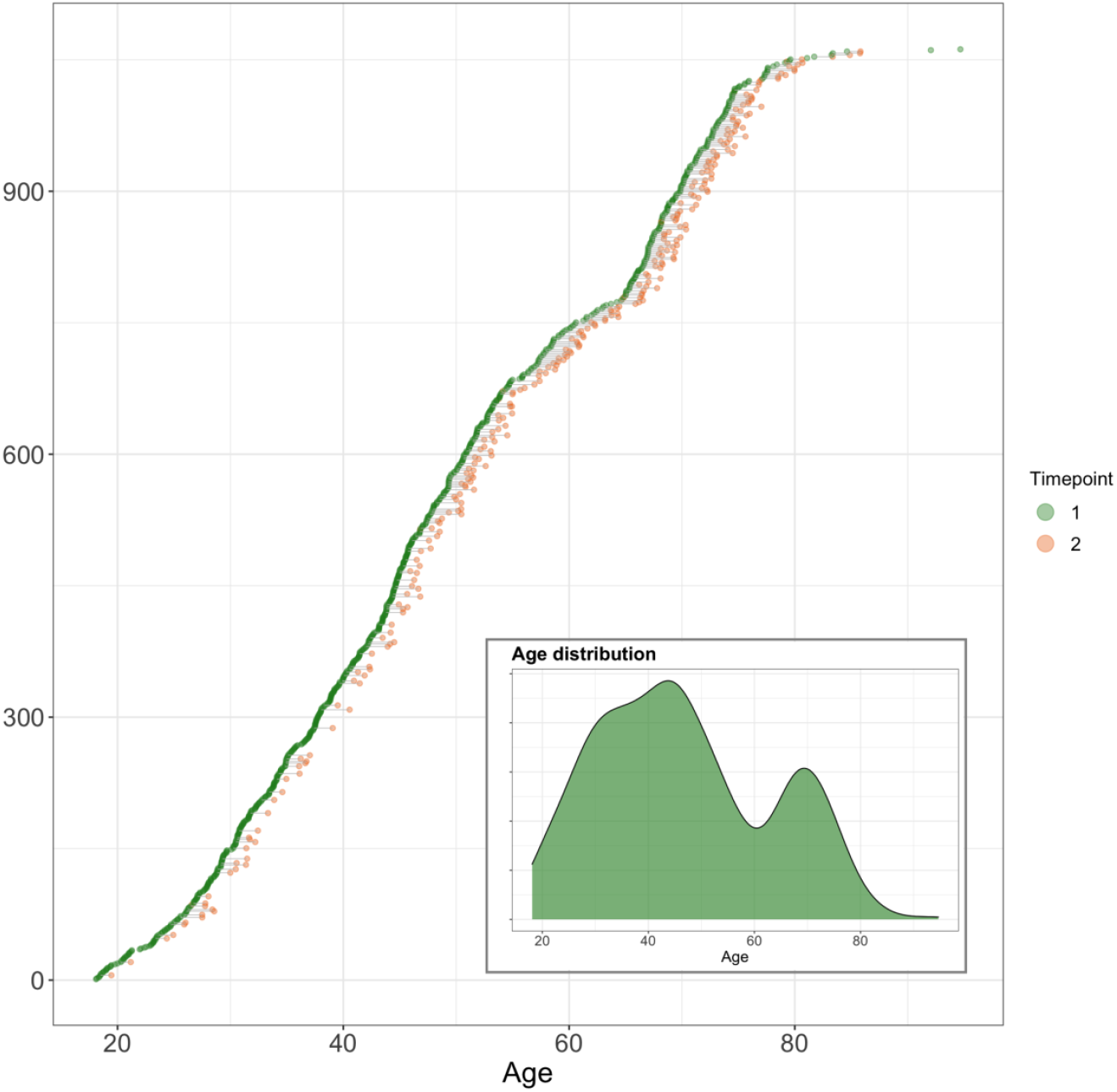
Available baseline and follow-up data. All participants are shown. Participants with data at baseline are visualised in green dots (N = 790). Of these participants, those with longitudinal measures of brain MRI are connected to corresponding timepoint two orange dots (N = 272). The y axis shows index which reorders data to sort by age at first timepoint. Subplot shows density of age distribution at baseline.

An independent training sample from the Cambridge Centre for Ageing and Neuroscience (Cam-CAN: http://www.mrc-cbu.cam.ac.uk/datasets/camcan/; Shafto et al., 2014; Taylor et al., 2017) was used for brain age prediction (section 2.6). After QC, MRI data from 622 participants were included (age range = 18–87, mean age ± standard deviation = 54.2 ± 18.4).

### 2.2. MRI acquisition

MRI was performed at Oslo University Hospital, Norway, on a GE Discovery MR750 3T scanner. Brain MRI was collected with a 32-channel head coil. T1-weighted data were acquired with a 32-channel head coil using a 3D inversion recovery prepared fast spoiled gradient recalled sequence (IR-FSPGR; BRAVO) with the following parameters: TR: 8.16 ms, TE: 3.18 ms, flip angle: 12°, voxel size: 1×1×1 mm^3^, FOV: 256 × 256 mm^2^, 188 sagittal slices, scan time: 4:43 min. DTI data were acquired with a spin echo planar imaging (EPI) sequence with the following parameters: repetition time (TR)/echo time (TE)/flip angle: 150 ms/83.1 ms/90°, FOV: 256 × 256 mm^2^, slice thickness: 2 mm, in-plane resolution: 2×2 mm, 60 non-coplanar directions (b = 1000 s/mm^2^) and 5 b = 0 volumes, scan time: 8:58 min. In addition, 7 b = 0 volumes with reversed phase-encoding direction were acquired.

Body MRI was performed with a single-slice 3D dual-echo LAVA Flex pulse sequence to acquire water and fat separated volumetric data covering head to knees. The following parameters were used: TE: minimum, flip angle: 10°, FOV: 50 x 50 mm, slice thickness: 5 mm, scan time: 2:32 min. For proton density fat fraction (PDFF) assessment in the liver, a single-slice 3D multi-echo IDEAL IQ pulse sequence was used with the following parameters: TE: minimum, flip angle: 3°, FOV: 45 x 45 mm^2^, slice thickness: 8 mm, scan time: 0:22 min.

The Cam-CAN training set participants were scanned on a 3T Siemens TIM Trio scanner with a 32-channel head-coil at Medical Research Council (UK) Cognition and Brain Sciences Unit (MRC-CBSU) in Cambridge, UK. High-resolution 3D T1-weighted data was collected using a magnetisation prepared rapid gradient echo (MPRAGE) sequence with the following parameters: TR: 2250 ms, TE: 2.99 ms, inversion time (TI): 900 ms, flip angle: 9°, FOV of 256 × 240 × 192 mm; voxel size = 1×1×1 mm, GRAPPA acceleration factor of 2, scan time 4:32 min (Dixon et al., 2014). DTI data was acquired using a twice-refocused spin echo sequence with the following parameters: TR: 9100 ms, TE: 104 ms, FOV: 192 × 192 mm, voxel size: 2 mm, 66 axial slices using 30 directions with b = 1000 s/mm^2^, 30 directions with b = 2000 s/mm^2^, and 3 b = 0 images (Dixon et al., 2014).

### 2.3. DTI processing and TBSS analysis

Processing steps for single-shell diffusion MRI data in the test set followed a previously described pipeline (Maximov et al., 2019), including noise correction (Veraart et al., 2016), Gibbs ringing correction (Kellner et al., 2016), corrections for susceptibility induced distortions, head movements and eddy current induced distortions using topup (http://fsl.fmrib.ox.ac.uk/fsl/fslwiki/topup) and eddy (http://fsl.fmrib.ox.ac.uk/fsl/fslwiki/eddy) (Andersson & Sotiropoulos, 2016). Isotropic smoothing was carried out with a Gaussian kernel of 1 mm^3^ implemented in the FSL function *fslmaths*. DTI metrics were estimated using *dtifit* in FSL and a weighted least squares algorithm. Processing steps for the training set followed a similar pipeline with the exception of the noise correction procedure.

Voxelwise analysis of the fractional anisotropy (FA) data was carried out using Tract-Based Spatial Statistics (TBSS) (Smith et al., 2006), as part of FSL (Smith et al., 2004). First, FA images were brain-extracted using BET (Smith, 2002) and aligned into a common space (FMRI58_FA template) using the nonlinear registration tool FNIRT (Andersson, Jenkinson, & Smith., 2007; Jenkinson et al., 2012), which uses a b-spline representation of the registration warp field (Rueckert et al., 1999). Next, the mean FA volume of all subjects was created and thinned to create a mean FA skeleton that represents the centres of all tracts common to the group. Each subject’s aligned FA data was then projected onto this skeleton. The mean FA skeleton was thresholded at FA > 0.2. This procedure was repeated in order to extract axial diffusivity (AD), mean diffusivity (MD), and radial diffusivity (RD). *fslmeants* was used to extract the mean skeleton and 20 regions of interest (ROI) based on a probabilistic white matter atlas (JHU) (Hua et al., 2008) for each metric. Including the mean skeleton values, 276 features per individual were derived in total.

### 2.4. FreeSurfer processing

T1-weighted MRI data were processed using FreeSurfer (Fischl, 2012) version 7.1.0 for the test set and version FreeSurfer version 5.3 for the training set. To extract reliable area, volume and thickness estimates, the test set including follow-up data were processed with the longitudinal stream (Reuter et al., 2012) in FreeSurfer. Specifically, an unbiased within-subject template space and image (Reuter & Fischl, 2011) is created using robust, inverse consistent registration (Reuter et al., 2010). Several processing steps, such as skull stripping, Talairach transforms, atlas registration as well as spherical surface maps and parcellations are then initialized with common information from the within-subject template, significantly increasing reliability and statistical power (Reuter et al., 2012). Due to the longitudinal stream in FreeSurfer influencing the thickness estimates, and subsequently having an impact on brain age prediction (Høgestøl et al., 2019), both cross-sectional and longitudinal data in the test set were processed with the longitudinal stream. All reconstructions were visually assessed and edited by trained research personnel. Cortical parcellation was performed using the Desikan-Killiany atlas (Desikan et al., 2006), and subcortical segmentation was performed using a probabilistic atlas (Fischl et al., 2002). 269 FreeSurfer based features were extracted in total, including global features for intracranial volume, total surface area, and whole cortex mean thickness, as well as the volume of subcortical structures.

### 2.5. Quality control (QC) procedure

A detailed description of the complete QC procedure for the final sample is available in (Beck et al., 2021b). Briefly, for DTI we derived various QC metrics (see Supplementary material; SI Table 1), including temporal signal-to-noise-ratio (tSNR) (Roalf et al., 2016) to flag data deemed to have unsatisfactory quality. For T1-weighted data, QC was carried out using the ENIGMA cortical QC protocol including three major steps: outlier detection, internal surface method, and external surface method. Following the removal of datasets with inadequate quality (n = 30), the separate T1 and DTI datasets were merged with BMI and WHR measures, leaving the final sample used for the study at N = 1062 datasets from 790 individuals, among which N = 286 had body MRI data available.

Body MRI QC was carried out using a multivariate outlier detection algorithm, where anomalies in the data are detected as observations that do not conform to an expected pattern to other items. Using the R package *mvoutlier* (Filzmoser et al., 2005), potential outliers were flagged using the Mahalanobis distance (SI Figures 1 and 2). Informed by an interactive plot using the *chisq*.*plot* function, manual outlier observations of each of these flagged values deemed none of them as true outliers, leading to no further removal from the initial 286 body MRI dataset.

### 2.6 Brain age prediction

We performed brain age prediction using T1-weighted and DTI data using XGBoost regression (https://xgboost.readthedocs.io/en/latest/python), which is based on a decision-tree ensemble algorithm used in several recent brain age prediction studies (Beck et al., 2021b; de Lange et al., 2019, 2020; Kaufmann et al., 2019; Richard et al., 2020). BAG was calculated using (predicted age-chronological age) for each of the models, providing T1 and DTI-based BAG values for each individual. The BAG estimates were residualised for age to account for age-bias (de Lange & Cole, 2020; Liang et al., 2019).

### 2.7. Adipose tissue measures

For body MRI measures of adipose tissue distribution, missing entries were identified (SI Figure 3) before being imputed using the *MICE* package (van Buuren & Groothuis-Oudshoorn, 2011) in R, where five imputations were carried out using the predictive mean matching method (package default). The distribution of the original and imputed data was inspected (SI Figure 4) and the imputed data were deemed as plausible values. Of the five imputations, the first was used for the remainder of the study.

To investigate the associations between the adipose tissue measures, hierarchical clustering of the variables was performed using *hclust*, part of the *stats* package in R (R Core Team, 2012), which uses the complete linkage method to form clusters. Five cluster groups were revealed. Principle component analysis (PCA) was performed using *prcomp*, part of the *stats* package in R (R Core Team, 2012), to visualise the variation present in the dataset. The PCA-derived scree plot (SI Figure 5) revealed that 79.4% of the variance was explained by the first two dimensions. SI Figure 6 provides a visualisation of the hierarchical clustering and SI Figure 7 provides a graph of PCA variables relatedness.

Informed by the cluster formations and PCA, left and right anterior and posterior thigh muscle and fat infiltration volumes were combined to form two average measures of thigh muscle volume and thigh muscle fat percentage. Moreover, raw data was converted to body composition features following calculations provided in (Linge et al., 2018). The final adipose tissue measures included liver fat, describing the PDFF in the liver; visceral adipose tissue index (VAT index), which is VAT normalised by height^2^, describing the intra-abdominal fat surrounding the organs; total adipose, which is the total abdominal fat (VAT and ASAT) normalised by height^2^; weight-to-muscle ratio (WMR), which is body weight divided by thigh muscle volume; fat ratio, which is the total abdominal fat divided by total abdominal fat and thigh muscle volume; and muscle fat infiltration (MFI). Figure 2 shows the body MRI for two participants. Figure 4 shows the associations between the adiposity measures in a network correlation graph, created using the *qgraph* (Epskamp et al., 2012) R package. For a correlation matrix of adiposity measures see SI Figure 8.

**Figure 2.**
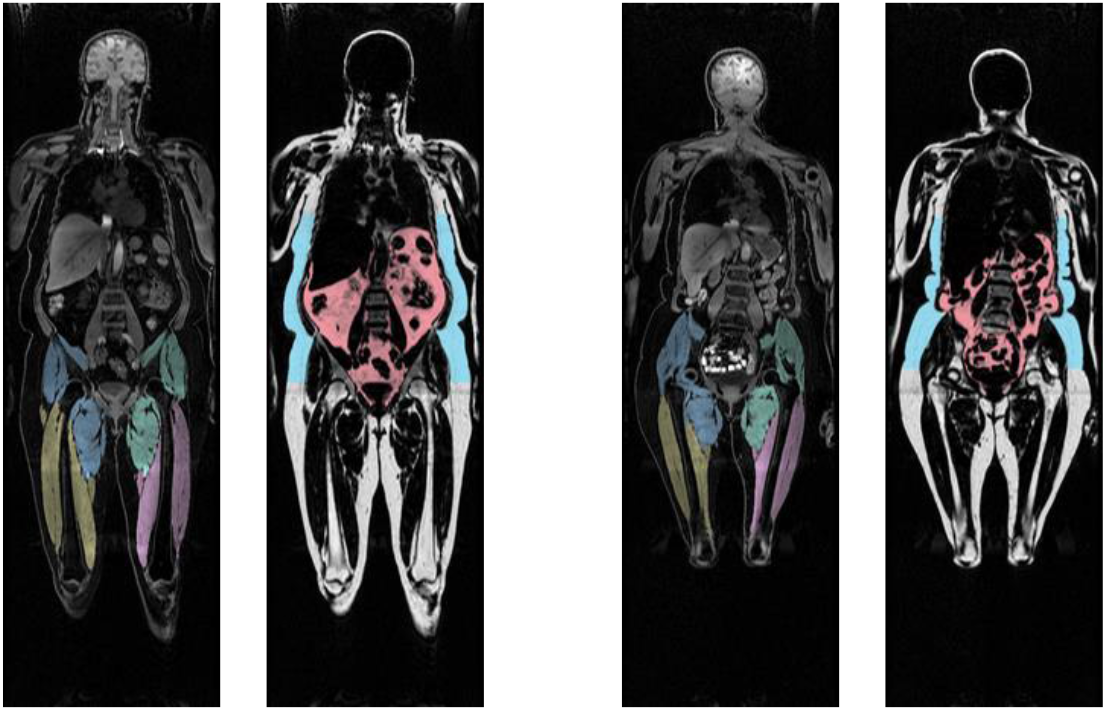
Body MRI. Showing two participants with a coronal slice from their MRI scan with VAT (pink) and ASAT (blue), and thigh muscle segmentations.

### 2.8. Statistical analysis

All statistical analyses were performed using R, version 3.6.0 (www.r-project.org/) (R Core Team, 2012). Bayesian multilevel models were carried out in *Stan* (Stan Development Team, 2019) using the *brms* (Bürkner, 2017, 2018) package. For descriptive purposes, we tested associations between each adiposity measure and age. Each adiposity measure was entered as the dependent variable while age was entered as the independent fixed effects along with sex and time.

To address the primary aim of the study, we tested for associations between tissue specific BAG and each adiposity measure. BAG (for T1 and DTI separately) was entered as the dependent variable with each adiposity measure separately entered as the independent fixed effects variable along with age, sex, and time.

To test our hypothesis that adiposity influences brain ageing we tested for associations between longitudinal changes in BAG and body MRI measures at follow-up using Bayesian multilevel models. Similarly, we tested for interactions between age and adiposity measures on BAG. For each of the models, timepoint and age were included in the models where appropriate, while sex was added to both models.

In order to prevent false positives and to regularize the estimated associations we defined a standard prior around zero with a standard deviation of 1 for all coefficients. For each coefficient of interest, we report the mean estimated value and its uncertainty measured by the 95% credible interval of the posterior distribution, and calculated Bayes factors (BF) using the Savage-Dickey method (Wagenmakers et al., 2010). For a pragmatic guide on Bayes factor (BF) interpretation, see SI Table 2.

## 3. Results

### 3.1. Brain age prediction

SI Table 3 summarises age prediction accuracy in the training and test sets. Briefly, the models revealed high accuracy, as previously reported (Beck et al., 2021b), with R^2^ = 0.72 and 0.73 for the T1 and DTI models, respectively.

### 3.2. Adiposity measures and brain age gap

#### 3.2.1. Descriptive statistics

Table 1 shows descriptive statistics, Figure 3 shows the distributions within women and men for each adiposity measure, and Figure 5 shows the correlations between adiposity measures for women and men.

**Figure 3.**
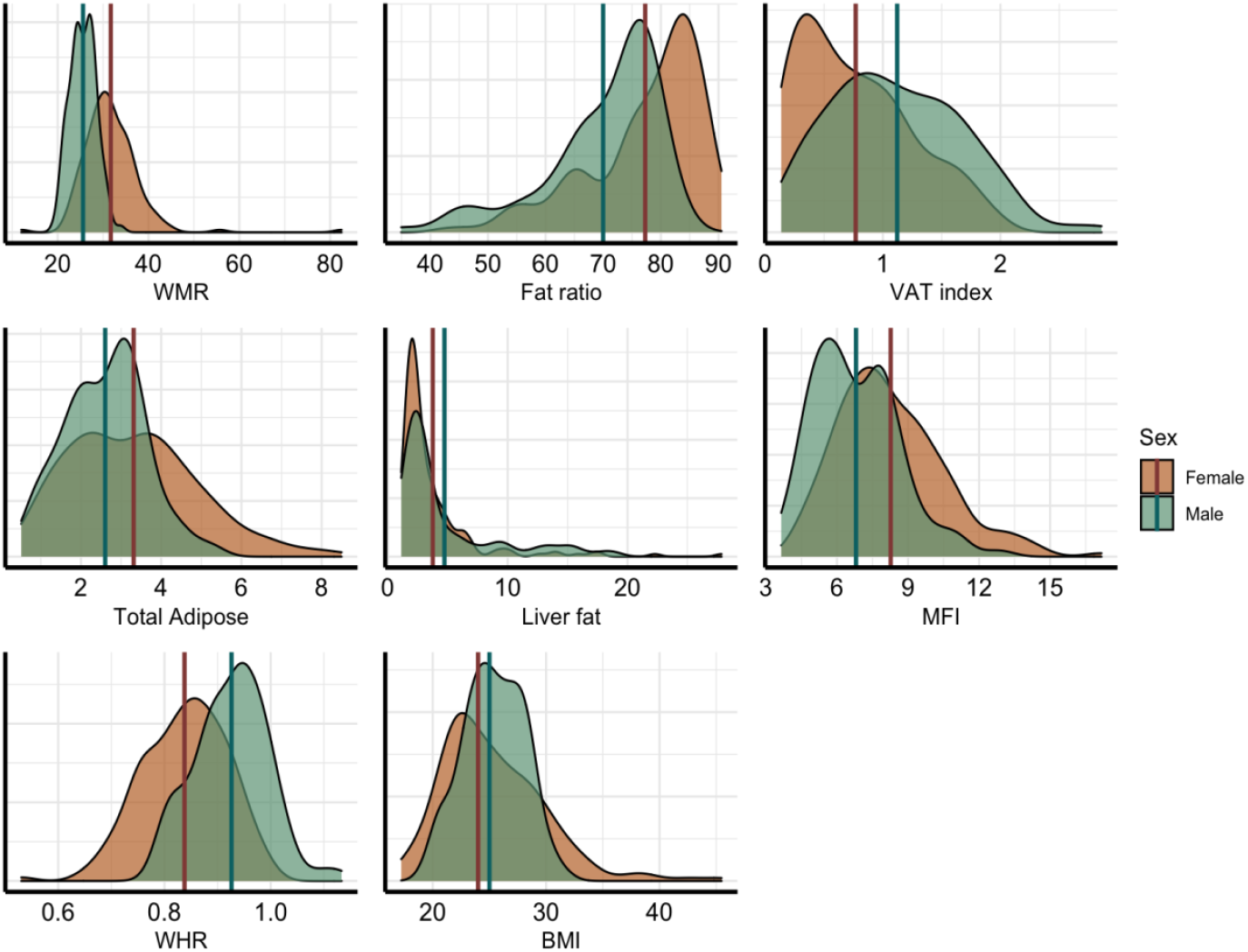
Distribution of the adiposity measures. Density plots for each variable, split by sex (male = green, female = orange). Vertical lines represent mean values for each sex.

**Figure 4.**
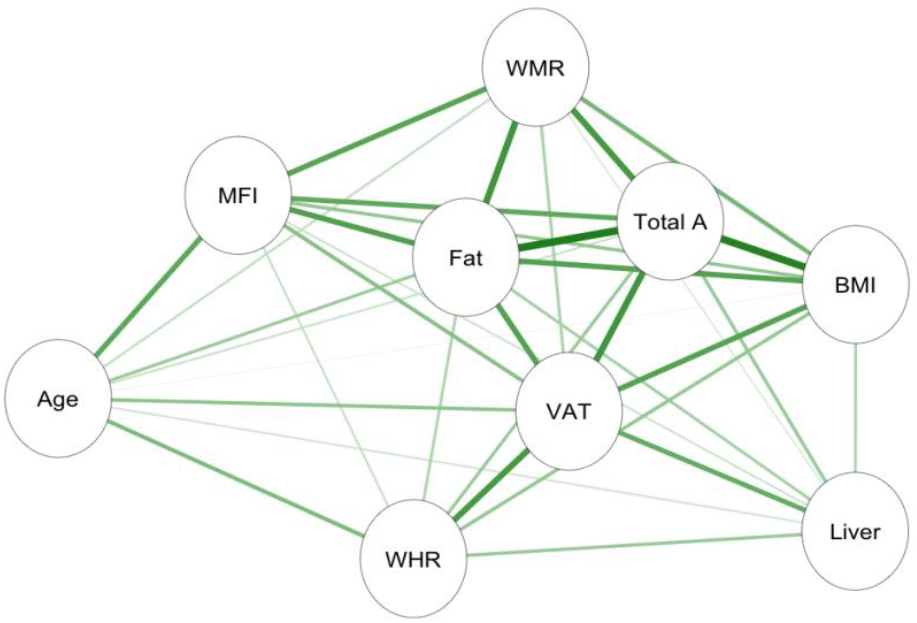
Associations between adiposity measures. Network correlation graph showing correlations between adiposity measures and age, where the green lines indicate positive associations, and orange lines (none present) indicate negative associations. Strength of association marked by thickness of each line, with the thickest line shown equating to r = 0.86. Abbreviations: MFI – muscle fat infiltration; Fat – fat ratio; WHR – waist-to-hip ratio; VAT – visceral abdominal tissue index; WMR – weight-to-muscle ratio; Total A – total adipose; BMI – body-mass index; Liver – liver fat.

**Figure 5.**
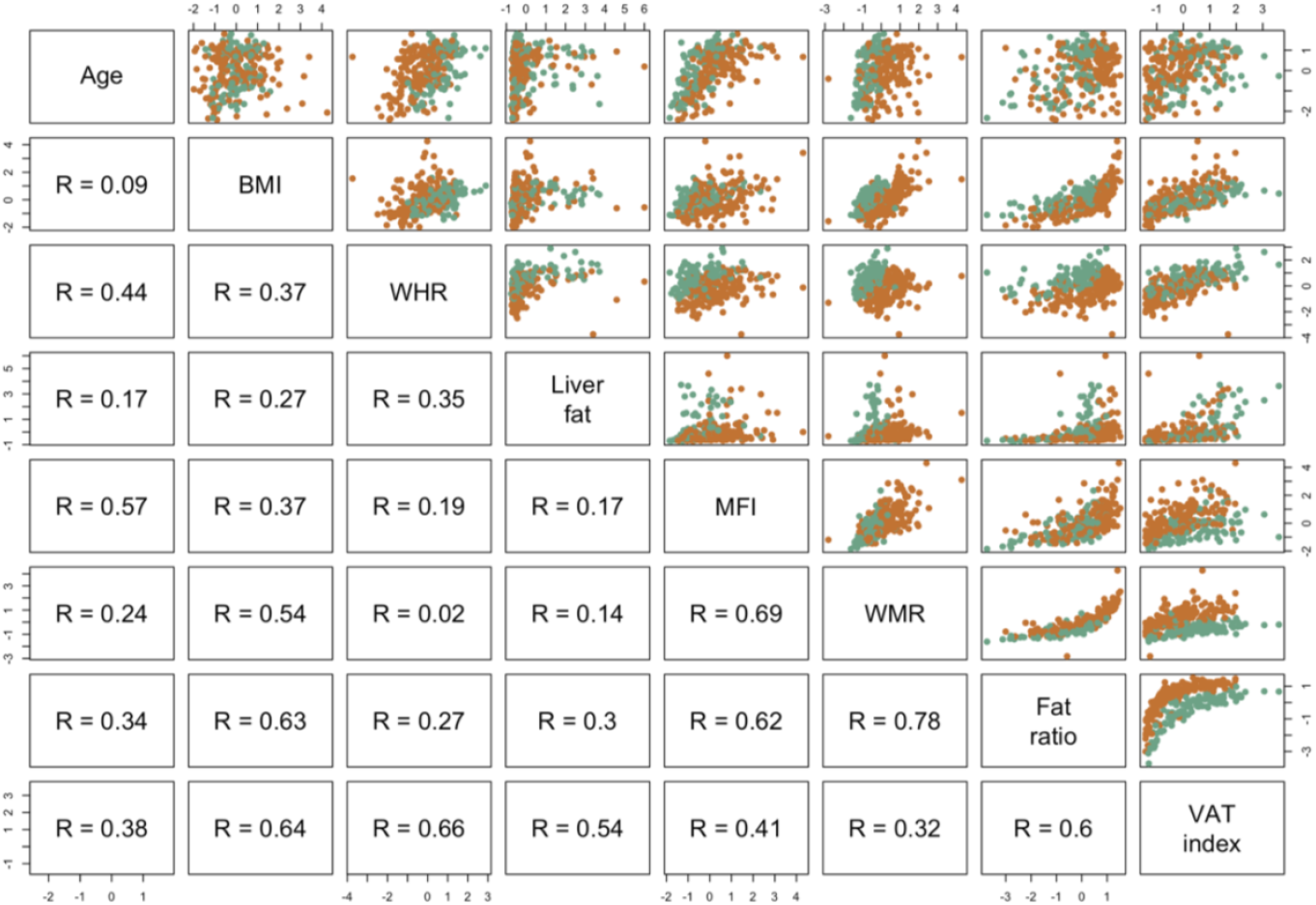
Associations between adiposity measures (and age) split by sex. Scatter plot matrix showing Pearson correlations between adiposity measures. Green points represent males, orange points represent females.

### 3.3. Bayesian multilevel models

#### 3.3.1. Descriptive results

For descriptive purposes, associations between each adiposity measure and age were tested. Results are summarised in the Supplementary Material, including visualisation of reported effects (SI Figures 9-10).

#### 3.3.2. Associations between BAG and adiposity measure

Figure 7 shows posterior distributions of the estimates of the coefficients reflecting the associations between each adiposity measure and BAGs, and Figure 8 shows credible intervals and evidence ratios. SI Figure 11 shows network correlation graph of correlations between adiposity measures and each BAG, and SI Table 5 provides summary statistics.

**Figure 6.**
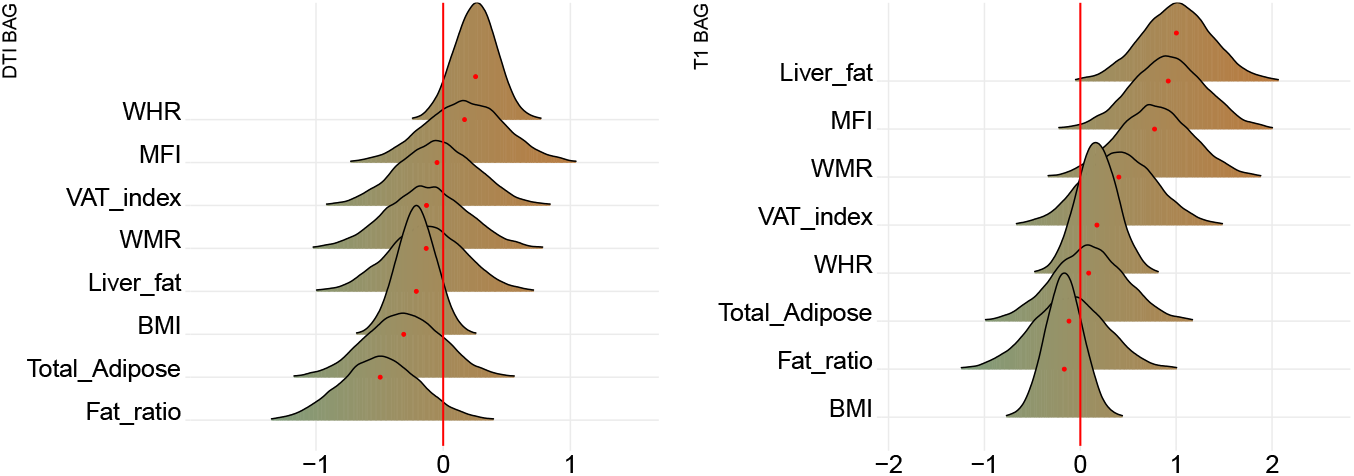
Associations between adiposity and BAG. The figure shows posterior distributions of the estimates of the standardised coefficient. Estimates for each variable on DTI BAG on the left and T1 BAG on the right. Colour scale follows direction evidence, with positive values indicating evidence in favour of an association and negative values evidence in favour of the null hypothesis. Width of distribution represents the uncertainty of the parameter estimates.

**Figure 7.**
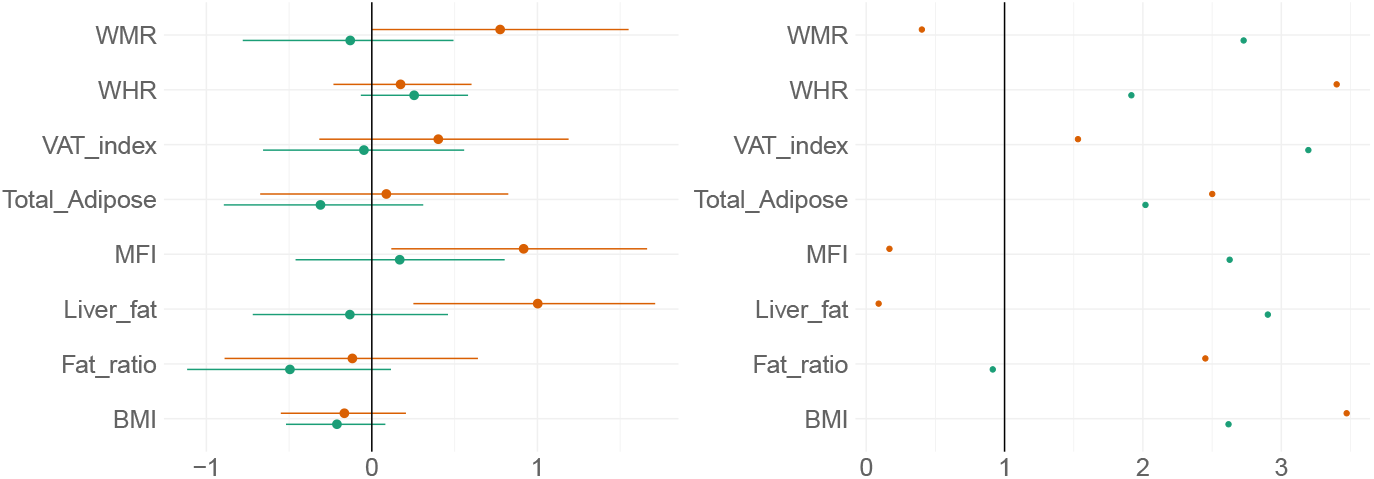
BAG and adiposity: Estimate credible intervals and evidence ratios. Left-side plot shows estimates with 95% credible intervals while the right-side figure shows likelihood of null where values above one indicate evidence in favour of the effect being null, and values below one indicate evidence in favour of an effect. T1 BAG associations are represented by orange points, and DTI BAG by green points.

**Figure 8.**
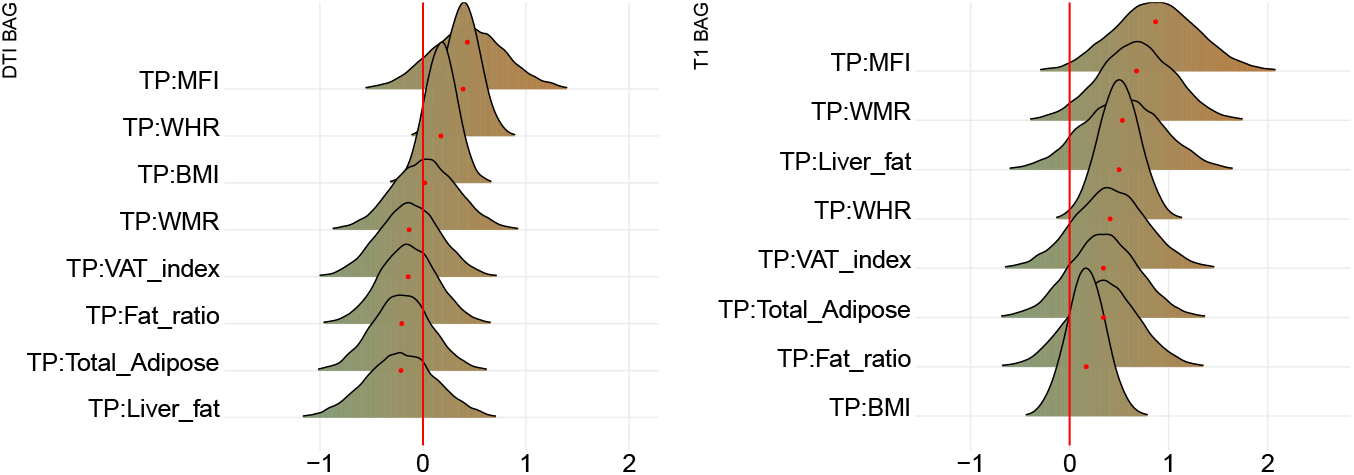
Interaction effects between adiposity and time on BAG. The figure shows posterior distributions of the estimates of the standardised coefficient. Estimates for the interaction effect of time and each adiposity measure on DTI BAG on the left and T1 BAG on the right.

The tests revealed anecdotal evidence in favour of an association with DTI BAG for fat ratio (BF = 0.91, *β = -*0.50), while moderate evidence in favour of no association was revealed between DTI BAG and VAT index (BF = 3.12, *β =* −0.05), liver fat (BF = 2.90, *β =* −0.13), and anecdotal evidence for WMR (BF = 2.73, *β = −*0.13), total adipose (BF = 2.02, *β =* −0.31), MFI (BF = 2.63, *β =* 0.17), WHR (BF = 1.92, *β =* 0.26), and BMI (BF = 2.62, *β = -* 0.21).

Strong evidence in favour of an association with T1 BAG was provided for liver fat (BF = 0.09, *β =* 1.0), with moderate evidence for MFI (BF = 0.17, *β =* 0.92), and anecdotal evidence for WMR (BF = 0.40, *β =* 0.77). The tests revealed moderate evidence in favour of no association between T1 BAG and WHR (BF = 3.40, *β =* 1.73), and BMI (BF = 3.47, *β =* - 0.17), and anecdotal evidence for fat ratio (BF = 2.5, *β =* −0.12), total adipose (BF = 2.50, *β =* 0.09), VAT index (BF = 1.53, *β =* 0.40).

#### 3.3.3. Interaction effects of time and adiposity measure on brain age gap

Figure 9 shows posterior distributions of the estimates of the coefficient for the interaction between time and each adiposity measure and DTI and T1 BAGs, and Figure 10 shows credible intervals and evidence ratios. For full table of results see SI Table 5.

**Figure 9.**
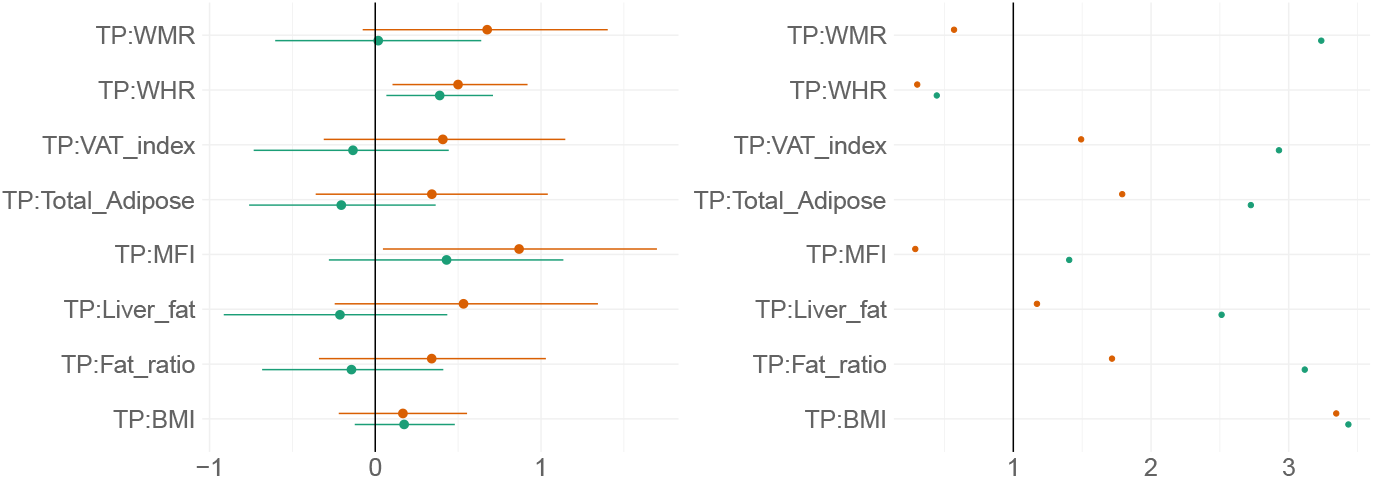
Interaction effects between adiposity and time on BAG: Estimate credible intervals and evidence ratios. Left-side plot shows estimates with 95% credible intervals while the right-side figure shows likelihood ratios. T1 BAG effects are represented by orange points, and DTI BAG by green points.

**Figure 10.**
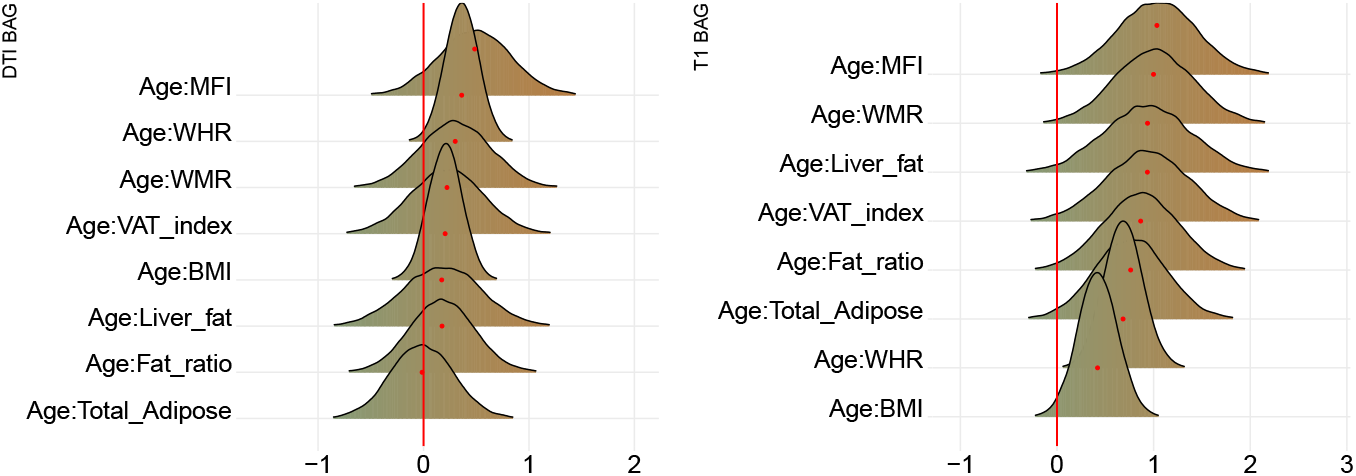
Interaction effects between adiposity and age on BAG. The figure shows posterior distributions of the estimates for the interaction effect between age and each variable on DTI BAG on the left and T1 BAG on the right.

For DTI BAG, the evidence supporting an interaction with time was anecdotal for WHR (BF = 0.44, *β =* 0.39) (SI Figure 12). The models revealed moderate evidence in favour of no interaction with time for WMR (BF = 3.24, *β =* 0.02), fat ratio (BF = 3.12, *β =* - 0.14), and BMI (BF = 3.43, *β =* 0.17), with anecdotal evidence for VAT index (BF = 2.93, *β =* −0.13), total adipose (BF = 2.73, *β =* −0.21), liver fat (BF = 2.51, *β =* −0.21), and MFI (BF = 1.41, *β =* 0.43).

For T1 BAG, the evidence supporting an interaction with time was moderate for WHR (BF = 0.30, *β =* 0.50) and MFI (BF = 0.29, *β =* 0.87), indicating faster pace of brain ageing among people with higher WHR and MFI (SI Figure 12). The models also revealed anecdotal evidence for WMR (BF = 0.57, *β =* 0.66). The evidence of no associations was anecdotal for fat ratio (BF = 1.72, *β =* 0.34), VAT index (BF = 1.49, *β =* 0.41), total adipose (BF = 1.79, *β =* 0.34), and liver fat (BF = 1.17, *β =* 0.53), and moderate for BMI (BF = 3.35, *β =* 0.17).

#### 3.3.4. Interaction effects of age and adiposity measure on brain age gap

Figure 11 shows posterior distributions of the estimates of the coefficient for the interaction between age and each adiposity measure and DTI and T1 BAGs, and Figure 12 shows credible intervals and evidence ratios. For full table of reported results see SI Table 5.

**Figure 11.**
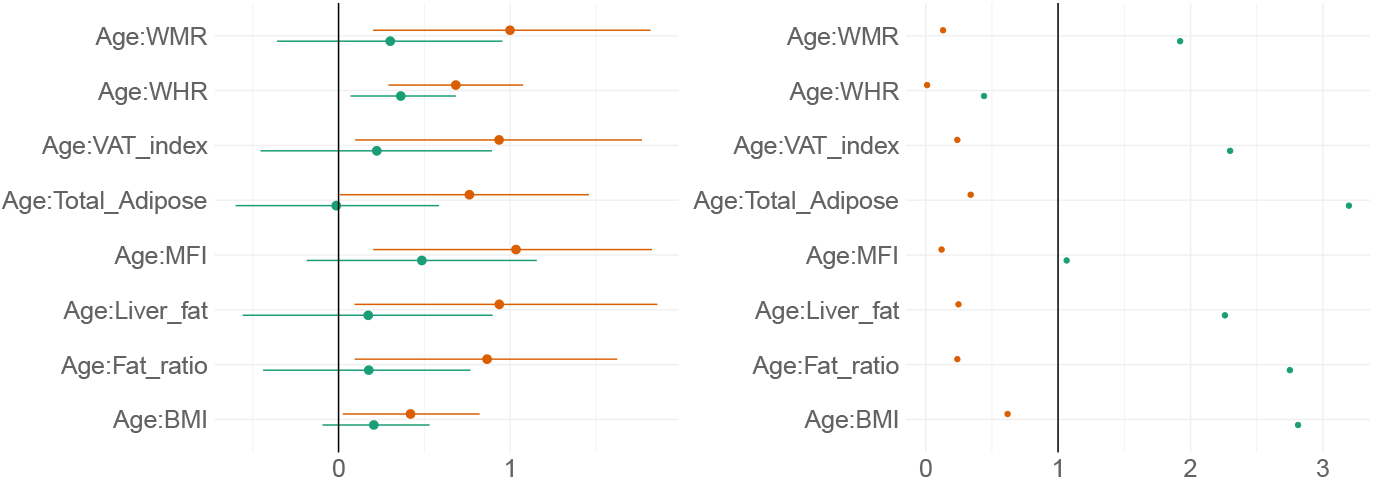
Interaction effects between adiposity and age on BAG: Estimate credible intervals and evidence ratios. Left-side plot shows estimates with 95% credible intervals while the right-side figure shows likelihood ratios. T1 BAG effects are represented by orange points, and DTI BAG by green points.

The analysis provided anecdotal evidence in support of an interaction effect with age on DTI BAG for WHR (BF = 0.44, *β =* 0.36). Anecdotal evidence was also provided in support of no interaction effect for WMR (BF = 1.92, *β =* 0.30), liver fat (BF = 2.26, *β =* 0.17), MFI (BF = 1.06, *β =* 0.49), fat ratio (BF = 2.75, *β =* 0.18), VAT index (BF = 2.29, *β =* 0.22), and BMI (BF = 2.81, *β =* 0.21), with moderate evidence for total adipose (BF = 3.20, *β =* −0.01).

The support of an interaction effect with age on T1 BAG was very strong for WHR (BF = 0.01, *β =* 0.68), and moderate for WMR (BF = 0.13, *β =* 1.00), fat ratio (BF = 0.24, *β =* 0.87), VAT index (BF = 0.24, *β =* 0.94), liver fat (BF = 0.25, *β =* 0.94), and MFI (BF = 0.12, *β =* 1.03), indicating these adiposity measures may be increasingly important predictors of BAG with increasing age. The models further indicated anecdotal evidence for total adipose (BF = 0.34, *β =* 0.76) and BMI (BF = 0.62, *β =* 0.42).

## 4. Discussion

Despite increasing evidence of shared mechanisms across several metabolic conditions and cardiovascular and neurodegenerative diseases, we are yet to fully understand the complex associations of adipose tissue and brain age. The current cross-sectional and longitudinal findings support that higher measures of adipose tissue – particularly higher liver fat and MFI – are associated with an older-appearing brain and faster brain ageing. Both overall brain age gap and the rates of change in brain age over time were associated with specific adipose tissue measures at follow-up, including thigh muscle fat infiltration, weight-to-muscle ratio, and liver fat.

### 4.1. Adiposity and brain age: cross-sectional analysis

Our findings demonstrated associations between liver fat, MFI, WMR and T1 BAG, indicating older-appearing brains in individuals with higher adipose tissue measures. DTI BAG associations were less common, with evidence supporting no associations with VAT index and WHR. These findings are in line with the hypothesis that body MRI adipose tissue measures are associated with ageing of the brain as indicated by brain MRI morphology measures. Moreover, the findings extend previous work linking adiposity measures, including links to grey matter volume (Debette & Markus, 2010; Gunstad et al., 2005; Jørgensen et al., 2017; Ward et al., 2005) and white matter microstructural properties based on diffusion MRI (Marks et al., 2011; Mueller et al., 2011; Stanek et al., 2011; Xu et al., 2013), with conflicting evidence for white matter volume (Friedman et al., 2014; Willette & Kapogiannis, 2015). The discrepancy may be due to methodological differences, e.g. between previous regional associations and our global brain age approach. Future research estimating regional brain age models trained with more advanced diffusion MRI features may offer improved sensitivity and specificity (Beck et al., 2021a).

MFI has previously been linked to metabolic risk factors (Therkelsen et al., 2013) and insulin resistance in obesity (Goodpaster et al., 2000). Higher liver fat has been found among diabetics (Bamberg et al., 2017; Linge et al., 2018) and prediabetics without previous cardiovascular conditions (Bamberg et al., 2017). Studies have also reported no significant association between CHD or cardiovascular events and elevated liver fat (Neeland et al., 2015). Moreover, while comparing patients with non-alcoholic fatty liver disease (NAFLD) to controls, Hagström et al., (2017) reported no significant difference in cardiovascular related death. Conflicting results however, have linked NAFLD with cardiovascular disease (Brea et al., 2017), suggesting a complex interplay between regional adipose tissue and metabolic health.

This complexity has recently been corroborated by findings of 62 genetic loci associated with both higher adiposity and lower cardiometabolic risk (Huang et al., 2021), possibly reflecting protective mechanisms. Although speculative, this heterogeneity is likely also reflected in connection to brain health, which for some individual genetic architectures may offer protection against brain pathology while simultaneously increasing the risk for obesity. Further studies exploiting larger sampler are needed to pursue this hypothesis and attempt to dissect the heterogeneity using genetic data (Huang et al., 2021) in combination with brain imaging.

### 4.2. Adiposity and brain ageing: longitudinal evidence

Our longitudinal analyses revealed that the rate of brain ageing across the study period was associated with adiposity measured at follow-up, with evidence for WHR from our anthropometric measures, and MFI and WMR from adipose tissue distribution measures. While evidence for WHR was present for both T1-weighted and DTI BAGs - which largely replicates a recent report from the same cohort (Beck et al., 2021b) - evidence for MFI and WMR were only observed for T1-weighted BAG, where greater adipose tissue was associated with increased rate of brain ageing. Conversely, evidence in favour of no interaction effect with time was found for BMI. This supports that BMI alone may not represent a clinically relevant proxy, as it fails to distinguish from lean mass while ignoring regional fat distribution, in particular the visceral components (Evans et al., 2012; Roberson et al., 2014).

While experimental evidence is needed to establish causality, our findings suggest that adipose tissue distribution affects brain ageing and may lead to older appearing brains in generally healthy individuals. These observations are in line with previous studies, both from clinical groups and population-based cohorts. Non-alcoholic fatty liver disease, for example, has been previously linked to smaller total brain volume (Weinstein et al., 2018) and cortical and cerebellar structures (Gurholt et al., 2020), while muscle fat infiltration has been negatively associated with cortical structures (Gurholt et al., 2020). While more research into adipose tissue distribution is warranted, the current results suggest that increased liver fat, muscle fat infiltration, and weight-to-muscle ratio may contribute to accelerate brain ageing.

### 4.3. Brain age and adiposity: interactions with age

For DTI BAG, our findings demonstrated an interaction effect with age and WHR, while T1 BAG interaction effects were present for age and WMR, fat ratio, total adipose, VAT index, liver fat, MFI, BMI, and WHR. The findings indicate that adiposity measures may be increasingly important predictors of BAG with increasing age. In contrast, we observed no evidence of interaction effects between age and BMI on DTI or T1 BAGs. Previous research has produced mixed results, with studies reporting an interaction of BMI and age on white matter volume but no interaction on cortical surface area or thickness (Ronan et al., 2016). Further research is warranted to elucidate the degree to which associations between adiposity and brain structure change over the course of the lifespan.

### 4.4. Strengths and limitations

The current study benefitted from a mixed cross-sectional and longitudinal design enabling brain changes to be tracked across time. The prediction models for brain age had high accuracy, and separate T1-weighted and DTI brain age gaps provided tissue-specific measures of brain age with potential to reveal specific associations with the included adiposity measures.

Several limitations should be considered when evaluating the findings. First, while the longitudinal brain MRI data represent a major strength, the follow-up sample size is relatively small, and the body MRI data was only collected at the follow-up examination. The subsequent loss of power is reflected in the width of body MRI posterior distributions, indicating a higher level of uncertainty compared to BMI and WHR, which had available longitudinal measures and a larger sample size. Next, although body composition measures based on MRI offer high accuracy in terms of fat and muscle distributions and are therefore a potentially valuable supplement to conventional anthropomorphic features, future studies including biological markers such as immune and inflammation assays and lipid measurements might provide even higher specificity and opportunities for further subtyping. Indeed, inflammation has demonstrated effects on brain function and structure (Rosano et al., 2011) and has been dubbed to have a central role in the obesity-brain connection (O’Brien et al., 2017). Moreover, including detailed assessments of dietary routines, alcohol intake, and physical exercise is vital in order to better understand the complex processes at play. For example, physical activity has been associated with lower brain age (Dunås et al., 2021; Sanders et al., 2021) and higher grey and white matter measures (Sexton et al., 2016), while excess alcohol intake is well documented in influencing liver and brain health (Agartz et al., 1999; Rehm et al., 2010). Lastly, the current sample is predominantly ethnic Scandinavian and Northern European, restricting our ability to generalise to wider populations, and future studies should aim to increase the diversity in the study population.

### 4.5. Conclusion

Combining body MRI and brain age prediction based on brain MRI allows for probing individual body composition profiles and brain patterns and trajectories which may confer risk for cardiometabolic disease and neurodegenerative disorders. More knowledge and further development of automated tools for individual phenotyping in this domain may inform public health priorities and interventions. With evidence of different adiposity subtypes being differentially linked with different brain phenotypes and cardiometabolic diseases, precision methods that look at fat distribution can potentially be more informative than conventional anthropomorphic measures. This in turn will provide a more effective tool for development of treatment strategies that focus on individual risk of metabolic disease, as well as disentangling the associations between body and brain health.

## Data Availability

The data and code used in the study will be freely available in a public repository (Open Science Framework) and eventually directly accessible through the OSF webpage.

https://osf.io/gws8x/

## 5. Acknowledgements

The study is supported by the Research Council of Norway (223273, 249795, 248238, 276082), the South-Eastern Norway Regional Health Authority (2014097, 2015044, 2015073, 2016083, 2018037, 2018076), the Norwegian ExtraFoundation for Health and Rehabilitation (2015/FO5146), KG Jebsen Stiftelsen, Swiss National Science Foundation (grant PZ00P3_193658), German Federal Ministry of Education and Research (BMBF, grant 01ZX1904A), ERA-Net Cofund through the ERA PerMed project ‘IMPLEMENT’ (Research Council of Norway – 298646), and the European Research Council under the European Union’s Horizon 2020 Research and Innovation program (ERC StG, Grant # 802998 and RIA Grant # 847776).

## References

Agartz, I., Momenan, R., Rawlings, R. R., Kerich, M. J., & Hommer, D. W. (1999). Hippocampal Volume in Patients With Alcohol Dependence. Archives of General Psychiatry, 56(4), 356–363. https://doi.org/10.1001/archpsyc.56.4.356

Andersson, J. L. R., & Sotiropoulos, S. N. (2016). An integrated approach to correction for off-resonance effects and subject movement in diffusion MR imaging. NeuroImage, 125, 1063–1078. https://doi.org/10.1016/j.neuroimage.2015.10.019

Anstey, K. J., Cherbuin, N., Budge, M., & Young, J. (2011). Body mass index in midlife and late-life as a risk factor for dementia: A meta-analysis of prospective studies: BMI and risk of dementia. Obesity Reviews, 12(5), e426–e437. https://doi.org/10.1111/j.1467-789X.2010.00825.x

Bahrami, S., Steen, N. E., Shadrin, A., O’Connell, K., Frei, O., Bettella, F., Wirgenes, K. V., Krull, F., Fan, C. C., Dale, A. M., Smeland, O. B., Djurovic, S., & Andreassen, O. A. (2020). Shared Genetic Loci Between Body Mass Index and Major Psychiatric Disorders: A Genome-wide Association Study. JAMA Psychiatry, 77(5), 503–512. https://doi.org/10.1001/jamapsychiatry.2019.4188

Bamberg, F., Hetterich, H., Rospleszcz, S., Lorbeer, R., Auweter, S. D., Schlett, C. L., Schafnitzel, A., Bayerl, C., Schindler, A., Saam, T., Müller-Peltzer, K., Sommer, W., Zitzelsberger, T., Machann, J., Ingrisch, M., Selder, S., Rathmann, W., Heier, M., Linkohr, B., … Peters, A. (2017). Subclinical Disease Burden as Assessed by Whole-Body MRI in Subjects With Prediabetes, Subjects With Diabetes, and Normal Control Subjects From the General Population: The KORA-MRI Study. Diabetes, 66(1), 158–169. https://doi.org/10.2337/db16-0630

Beck, D., de Lange, A.-M. G., Pedersen, M. L., Aln, D., Voldsbekk, I., Richard, G., Sanders, A.-M., Dørum, E. S., Kolskår, K. K., Høgestøl, E. A., Steen, N. E., Andreassen, O. A., Nordvik, J. E., Kaufmann, T., & Westlye, L. T. (n.d.). Cardiometabolic risk factors associated with brain age and accelerate brain ageing. 47.

Beck, D., de Lange, A.-M., Maximov, I. I., Richard, G., Andreassen, O. A., Nordvik, J. E., & Westlye, L. T. (2021a). White matter microstructure across the adult lifespan: A mixed longitudinal and cross-sectional study using advanced diffusion models and brain-age prediction. NeuroImage, 224, 117441. https://doi.org/10.1016/j.neuroimage.2020.117441

Beck, D., Lange, A.-M. G. de, Pedersen, M. L., Alnæs, D., Maximov, I. I., Voldsbekk, I., Richard, G., Sanders, A.-M., Ulrichsen, K. M., Dørum, E. S., Kolskår, K. K., Høgestøl, E. A., Steen, N. E., Djurovic, S., Andreassen, O. A., Nordvik, J. E., Kaufmann, T., & Westlye, L. T. (2021b). Cardiometabolic risk factors associated with brain age and accelerate brain ageing. MedRxiv, 2021.02.25.21252272. https://doi.org/10.1101/2021.02.25.21252272

Bhupathiraju, S. N., & Hu, F. B. (2016). Epidemiology of Obesity and Diabetes and Their Cardiovascular Complications. Circulation Research, 118(11), 1723–1735. https://doi.org/10.1161/CIRCRESAHA.115.306825

Brea, Á., Pintó, X., Ascaso, J. F., Blasco, M., Díaz, Á., González-Santos, P., Hernández Mijares, A., Mantilla, T., Millán, J., & Pedro-Botet, J. (2017). Nonalcoholic fatty liver disease, association with cardiovascular disease and treatment. (I). Nonalcoholic fatty liver disease and its association with cardiovascular disease. Clínica e Investigación En Arteriosclerosis (English Edition), 29(3), 141–148. https://doi.org/10.1016/j.artere.2016.06.001

Bürkner, P.-C. (2017). brms: An R Package for Bayesian Multilevel Models Using Stan. Journal of Statistical Software, 80(1). https://doi.org/10.18637/jss.v080.i01

Bürkner, P.-C. (2018). Advanced Bayesian Multilevel Modeling with the R Package brms. The R Journal, 10(1), 395. https://doi.org/10.32614/RJ-2018-017

Cole, J. H. (2020). Multimodality neuroimaging brain-age in UK biobank: Relationship to biomedical, lifestyle, and cognitive factors. Neurobiology of Aging, 92, 34–42. https://doi.org/10.1016/j.neurobiolaging.2020.03.014

Cole, J. H., Poudel, R. P. K., Tsagkrasoulis, D., Caan, M. W. A., Steves, C., Spector, T. D., & Montana, G. (2017). Predicting brain age with deep learning from raw imaging data results in a reliable and heritable biomarker. NeuroImage, 163, 115–124. https://doi.org/10.1016/j.neuroimage.2017.07.059

de Lange, A.-M., Barth, C., Kaufmann, T., Maximov, I. I., van der Meer, D., Agartz, I., & Westlye, L. T. (2019). Cumulative estrogen exposure, APOE genotype, and women’s brain aging—A population-based neuroimaging study [Preprint]. Neuroscience. https://doi.org/10.1101/826123

de Lange, A.-M. G., Anatürk, M., Kaufmann, T., Cole, J. H., Griffanti, L., Zsoldos, E., Jensen, D., Suri, S., Filippini, N., Singh-Manoux, A., Kivimäki, M., Westlye, L. T., & Ebmeier, K. P. (2020). Multimodal brain-age prediction and cardiovascular risk: The Whitehall II MRI sub-study [Preprint]. Neuroscience. https://doi.org/10.1101/2020.01.28.923094

de Lange, A.-M. G., & Cole, J. H. (2020). Commentary: Correction procedures in brain-age prediction. NeuroImage: Clinical, 26, 102229. https://doi.org/10.1016/j.nicl.2020.102229

Debette, S., & Markus, H. S. (2010). The clinical importance of white matter hyperintensities on brain magnetic resonance imaging: Systematic review and meta-analysis. BMJ, 341(jul26 1), c3666–c3666. https://doi.org/10.1136/bmj.c3666

Desikan, R. S., Ségonne, F., Fischl, B., Quinn, B. T., Dickerson, B. C., Blacker, D., Buckner, R. L., Dale, A. M., Maguire, R. P., Hyman, B. T., Albert, M. S., & Killiany, R. J. (2006). An automated labeling system for subdividing the human cerebral cortex on MRI scans into gyral based regions of interest. NeuroImage, 31(3), 968–980. https://doi.org/10.1016/j.neuroimage.2006.01.021

Ditmars, H. L., Logue, M. W., Toomey, R., McKenzie, R. E., Franz, C. E., Panizzon, M. S., Reynolds, C. A., Cuthbert, K. N., Vandiver, R., Gustavson, D. E., Eglit, G. M. L., Elman, J. A., Sanderson-Cimino, M., Williams, M. E., Andreassen, O. A., Dale, A. M., Eyler, L. T., Fennema-Notestine, C., Gillespie, N. A., … Lyons, M. J. (2021). Associations between depression and cardiometabolic health: A 27-year longitudinal study. Psychological Medicine, 1–11. https://doi.org/10.1017/S003329172000505X

Dixon, M., Taylor, J. R., Rowe, J. B., Cusack, R., Calder, A. J., Marslen-Wilson, W. D., Duncan, J., Dalgleish, T., Henson, R. N., Brayne, C., & Matthews, F. E. (2014). The Cambridge Centre for Ageing and Neuroscience (Cam-CAN) study protocol: A cross-sectional, lifespan, multidisciplinary examination of healthy cognitive ageing. BMC Neurology, 14(1), 204. https://doi.org/10.1186/s12883-014-0204-1

Dunås, T., Wåhlin, A., Nyberg, L., & Boraxbekk, C.-J. (2021). Multimodal Image Analysis of Apparent Brain Age Identifies Physical Fitness as Predictor of Brain Maintenance. Cerebral Cortex, bhab019. https://doi.org/10.1093/cercor/bhab019

Epskamp, S., Cramer, A., Waldorp, L., Schmittmann, V., & Borsboom, D. (2012). qgraph: Network Visualizations of Relationships in Psychometric Data. Journal of Statistical Software, 48. https://doi.org/10.18637/jss.v048.i04

Evans, P. D., McIntyre, N. J., Fluck, R. J., McIntyre, C. W., & Taal, M. W. (2012). Anthropomorphic Measurements That Include Central Fat Distribution Are More Closely Related with Key Risk Factors than BMI in CKD Stage 3. PLoS ONE, 7(4). https://doi.org/10.1371/journal.pone.0034699

Filzmoser, P., Garrett, R. G., & Reimann, C. (2005). Multivariate outlier detection in exploration geochemistry$. 9.

Fischl, B., Salat, D. H., Busa, E., Albert, M., Dieterich, M., Haselgrove, C., van der Kouwe, A., Killiany, R., Kennedy, D., Klaveness, S., Montillo, A., Makris, N., Rosen, B., & Dale, A. M. (2002). Whole Brain Segmentation. Neuron, 33(3), 341–355. https://doi.org/10.1016/S0896-6273(02)00569-X

Franke, K., Ristow, M., & Gaser, C. (2014). Gender-specific impact of personal health parameters on individual brain aging in cognitively unimpaired elderly subjects. Frontiers in Aging Neuroscience, 6. https://doi.org/10.3389/fnagi.2014.00094

Friedman, J. I., Tang, C. Y., de Haas, H. J., Changchien, L., Goliasch, G., Dabas, P., Wang, V., Fayad, Z. A., Fuster, V., & Narula, J. (2014). Brain imaging changes associated with risk factors for cardiovascular and cerebrovascular disease in asymptomatic patients. JACC. Cardiovascular Imaging, 7(10), 1039–1053. https://doi.org/10.1016/j.jcmg.2014.06.014

Goodpaster, B. H., Thaete, F. L., & Kelley, D. E. (2000). Thigh adipose tissue distribution is associated with insulin resistance in obesity and in type 2 diabetes mellitus. The American Journal of Clinical Nutrition, 71(4), 885–892. https://doi.org/10.1093/ajcn/71.4.885

Gunstad, J., Cohen, R. A., Tate, D. F., Paul, R. H., Poppas, A., Hoth, K., Macgregor, K. L., & Jefferson, A. L. (2005). Blood pressure variability and white matter hyperintensities in older adults with cardiovascular disease. Blood Pressure, 14(6), 353–358. https://doi.org/10.1080/08037050500364117

Gurholt, T. P., Kaufmann, T., Frei, O., Alnæs, D., Haukvik, U. K., van der Meer, D., Moberget, T., O’Connell, K. S., Leinhard, O. D., Linge, J., Simon, R., Smeland, O. B., Sønderby, I. E., Winterton, A., Steen, N. E., Westlye, L. T., & Andreassen, O. A. (2020). Population-based body-brain mapping links brain morphology and body composition [Preprint]. Neuroscience. https://doi.org/10.1101/2020.02.29.970095

Hagström, H., Nasr, P., Ekstedt, M., Hammar, U., Stål, P., Hultcrantz, R., & Kechagias, S. (2017). Fibrosis stage but not NASH predicts mortality and time to development of severe liver disease in biopsy-proven NAFLD. Journal of Hepatology, 67(6), 1265–1273. https://doi.org/10.1016/j.jhep.2017.07.027

Han, L. K. M., Dinga, R., Hahn, T., Ching, C. R. K., Eyler, L. T., Aftanas, L., Aghajani, M., Aleman, A., Baune, B. T., Berger, K., Brak, I., Filho, G. B., Carballedo, A., Connolly, C. G., Couvy-Duchesne, B., Cullen, K. R., Dannlowski, U., Davey, C. G., Dima, D., … Schmaal, L. (2020). Brain aging in major depressive disorder: Results from the ENIGMA major depressive disorder working group. Molecular Psychiatry. https://doi.org/10.1038/s41380-020-0754-0

Høgestøl, E. A., Kaufmann, T., Nygaard, G. O., Beyer, M. K., Sowa, P., Nordvik, J. E., Kolskår, K., Richard, G., Andreassen, O. A., Harbo, H. F., & Westlye, L. T. (2019). Cross-Sectional and Longitudinal MRI Brain Scans Reveal Accelerated Brain Aging in Multiple Sclerosis. Frontiers in Neurology, 10, 450. https://doi.org/10.3389/fneur.2019.00450

Hua, K., Zhang, J., Wakana, S., Jiang, H., Li, X., Reich, D. S., Calabresi, P. A., Pekar, J. J., van Zijl, P. C. M., & Mori, S. (2008). Tract probability maps in stereotaxic spaces: Analyses of white matter anatomy and tract-specific quantification. NeuroImage, 39(1), 336–347. https://doi.org/10.1016/j.neuroimage.2007.07.053

Huang, L. O., Rauch, A., Mazzaferro, E., Preuss, M., Carobbio, S., Bayrak, C. S., Chami, N., Wang, Z., Schick, U. M., Yang, N., Itan, Y., Vidal-Puig, A., den Hoed, M., Mandrup, S., Kilpeläinen, T. O., & Loos, R. J. F. (2021). Genome-wide discovery of genetic loci that uncouple excess adiposity from its comorbidities. Nature Metabolism, 3(2), 228–243. https://doi.org/10.1038/s42255-021-00346-2

Iacobini, C., Pugliese, G., Blasetti Fantauzzi, C., Federici, M., & Menini, S. (2019). Metabolically healthy versus metabolically unhealthy obesity. Metabolism, 92, 51–60. https://doi.org/10.1016/j.metabol.2018.11.009

Jenkinson, M., Beckmann, C. F., Behrens, T. E. J., Woolrich, M. W., & Smith, S. M. (2012). FSL. NeuroImage, 62(2), 782–790. https://doi.org/10.1016/j.neuroimage.2011.09.015

Jørgensen, K. N., Nesvåg, R., Nerland, S., Mørch-Johnsen, L., Westlye, L. T., Lange, E. H., Haukvik, U. K., Hartberg, C. B., Melle, I., Andreassen, O. A., & Agartz, I. (2017). Brain volume change in first-episode psychosis: An effect of antipsychotic medication independent of BMI change. Acta Psychiatrica Scandinavica, 135(2), 117–126. https://doi.org/10.1111/acps.12677

Kaufmann, T., Meer, D. van der, Doan, N. T., Schwarz, E., Lund, M. J., Agartz, I., Alnæs, D., Barch, D. M., Baur-Streubel, R., Bertolino, A., Bettella, F., Beyer, M. K., Bøen, E., Borgwardt, S., Brandt, C. L., Buitelaar, J., Celius, E. G., Cervenka, S., Conzelmann, A., … Westlye, L. T. (2019). Common brain disorders are associated with heritable patterns of apparent aging of the brain. Nature Neuroscience, 22(10), 1617–1623. https://doi.org/10.1038/s41593-019-0471-7

Kaufmann, T., van der Meer, D., Doan, N. T., Schwarz, E., Lund, M. J., Agartz, I., Alnæs, D., Barch, D. M., Baur-Streubel, R., Bertolino, A., Bettella, F., Beyer, M. K., Bøen, E., Borgwardt, S., Brandt, C. L., Buitelaar, J., Celius, E. G., Cervenka, S., Conzelmann, A., … Westlye, L. T. (2019). Common brain disorders are associated with heritable patterns of apparent aging of the brain. Nature Neuroscience, 22(10), 1617–1623. https://doi.org/10.1038/s41593-019-0471-7

Kellner, E., Dhital, B., Kiselev, V. G., & Reisert, M. (2016). Gibbs-ringing artifact removal based on local subvoxel-shifts. Magnetic Resonance in Medicine, 76(5), 1574–1581. https://doi.org/10.1002/mrm.26054

Kolbeinsson, A., Filippi, S., Panagakis, Y., Matthews, P. M., Elliott, P., Dehghan, A., & Tzoulaki, I. (2020). Accelerated MRI-predicted brain ageing and its associations with cardiometabolic and brain disorders. Scientific Reports, 10(1), 19940. https://doi.org/10.1038/s41598-020-76518-z

Kolenic, M., Franke, K., Hlinka, J., Matejka, M., Capkova, J., Pausova, Z., Uher, R., Alda, M., Spaniel, F., & Hajek, T. (2018). Obesity, dyslipidemia and brain age in first-episode psychosis. Journal of Psychiatric Research, 99, 151–158. https://doi.org/10.1016/j.jpsychires.2018.02.012

Liang, H., Zhang, F., & Niu, X. (2019). Investigating systematic bias in brain age estimation with application to post-traumatic stress disorders. Human Brain Mapping, hbm.24588. https://doi.org/10.1002/hbm.24588

Linge, J., Borga, M., West, J., Tuthill, T., Miller, M. R., Dumitriu, A., Thomas, E. L., Romu, T., Tunón, P., Bell, J. D., & Dahlqvist Leinhard, O. (2018). Body Composition Profiling in the UK Biobank Imaging Study: Body Composition Profiling in UK Biobank. Obesity, 26(11), 1785–1795. https://doi.org/10.1002/oby.22210

Linge, J., Ekstedt, M., & Dahlqvist Leinhard, O. (2021). Adverse muscle composition is linked to poor functional performance and metabolic comorbidities in NAFLD. JHEP Reports, 3(1), 100197. https://doi.org/10.1016/j.jhepr.2020.100197

Linge, J., Heymsfield, S. B., & Dahlqvist Leinhard, O. (2020). On the Definition of Sarcopenia in the Presence of Aging and Obesity—Initial Results from UK Biobank. The Journals of Gerontology: Series A, 75(7), 1309–1316. https://doi.org/10.1093/gerona/glz229

Linge, J., Whitcher, B., Borga, M., & Leinhard, O. D. (2019). Sub-phenotyping Metabolic Disorders Using Body Composition: An Individualized, Nonparametric Approach Utilizing Large Data Sets. Obesity, 27(7), 1190–1199. https://doi.org/10.1002/oby.22510

Luppino, F. S., de Wit, L. M., Bouvy, P. F., Stijnen, T., Cuijpers, P., Penninx, B. W. J. H., & Zitman, F. G. (2010). Overweight, obesity, and depression: A systematic review and meta-analysis of longitudinal studies. Archives of General Psychiatry, 67(3), 220–229. https://doi.org/10.1001/archgenpsychiatry.2010.2

Marks, B. L., Katz, L. M., Styner, M., & Smith, J. K. (2011). Aerobic fitness and obesity: Relationship to cerebral white matter integrity in the brain of active and sedentary older adults. British Journal of Sports Medicine, 45(15), 1208–1215. https://doi.org/10.1136/bjsm.2009.068114

Maximov, I. I., Alnæs, D., & Westlye, L. T. (2019). Towards an optimised processing pipeline for diffusion magnetic resonance imaging data: Effects of artefact corrections on diffusion metrics and their age associations in UK Biobank. Human Brain Mapping, 40(14), 4146–4162. https://doi.org/10.1002/hbm.24691

Mueller, K., Anwander, A., Möller, H. E., Horstmann, A., Lepsien, J., Busse, F., Mohammadi, S., Schroeter, M. L., Stumvoll, M., Villringer, A., & Pleger, B. (2011). Sex-Dependent Influences of Obesity on Cerebral White Matter Investigated by Diffusion-Tensor Imaging. PLOS ONE, 6(4), e18544. https://doi.org/10.1371/journal.pone.0018544

Mulugeta, A., Lumsden, A., & Hyppönen, E. (2021). Unlocking the causal link of metabolically different adiposity subtypes with brain volumes and the risks of dementia and stroke: A Mendelian randomization study. Neurobiology of Aging, S0197458021000658. https://doi.org/10.1016/j.neurobiolaging.2021.02.010

Neeland, I. J., Turer, A. T., Ayers, C. R., Berry, J. D., Rohatgi, A., Das, S. R., Khera, A., Vega, G. L., McGuire, D. K., Grundy, S. M., & de Lemos, J. A. (2015). Body Fat Distribution and Incident Cardiovascular Disease in Obese Adults. Journal of the American College of Cardiology, 65(19), 2150–2151. https://doi.org/10.1016/j.jacc.2015.01.061

O’Brien, P. D., Hinder, L. M., Callaghan, B. C., & Feldman, E. L. (2017). Neurological consequences of obesity. The Lancet Neurology, 16(6), 465–477. https://doi.org/10.1016/S1474-4422(17)30084-4

Pannacciulli, N., Del Parigi, A., Chen, K., Le, D. S. N. T., Reiman, E. M., & Tataranni, P. A. (2006). Brain abnormalities in human obesity: A voxel-based morphometric study. NeuroImage, 31(4), 1419–1425. https://doi.org/10.1016/j.neuroimage.2006.01.047

Pardoe, H. R., Cole, J. H., Blackmon, K., Thesen, T., & Kuzniecky, R. (2017). Structural brain changes in medically refractory focal epilepsy resemble premature brain aging. Epilepsy Research, 133, 28–32. https://doi.org/10.1016/j.eplepsyres.2017.03.007

Perry, B. I., Stochl, J., Upthegrove, R., Zammit, S., Wareham, N., Langenberg, C., Winpenny, E., Dunger, D., Jones, P. B., & Khandaker, G. M. (2021). Longitudinal Trends in Childhood Insulin Levels and Body Mass Index and Associations With Risks of Psychosis and Depression in Young Adults. JAMA Psychiatry. https://doi.org/10.1001/jamapsychiatry.2020.4180

Qiu, C., & Fratiglioni, L. (2015). A major role for cardiovascular burden in age-related cognitive decline. Nature Reviews Cardiology, 12(5), 267–277. https://doi.org/10.1038/nrcardio.2014.223

Quintana, D. S., Dieset, I., Elvsåshagen, T., Westlye, L. T., & Andreassen, O. A. (2017). Oxytocin system dysfunction as a common mechanism underlying metabolic syndrome and psychiatric symptoms in schizophrenia and bipolar disorders. Frontiers in Neuroendocrinology, 45, 1–10. https://doi.org/10.1016/j.yfrne.2016.12.004

Rajan, T., & Menon, V. (2017). Psychiatric disorders and obesity: A review of association studies. Journal of Postgraduate Medicine, 63(3), 182–190. https://doi.org/10.4103/jpgm.JPGM_712_16

Raji, C. A., Ho, A. J., Parikshak, N. N., Becker, J. T., Lopez, O. L., Kuller, L. H., Hua, X., Leow, A. D., Toga, A. W., & Thompson, P. M. (2010). Brain structure and obesity. Human Brain Mapping, 31(3), 353–364. https://doi.org/10.1002/hbm.20870

Rapuano, K. M., Laurent, J. S., Hagler, D. J., Hatton, S. N., Thompson, W. K., Jernigan, T. L., Dale, A. M., Casey, B. J., & Watts, R. (2020). Nucleus accumbens cytoarchitecture predicts weight gain in children. Proceedings of the National Academy of Sciences, 117(43), 26977–26984. https://doi.org/10.1073/pnas.2007918117

Rehm, J., Taylor, B., Mohapatra, S., Irving, H., Baliunas, D., Patra, J., & Roerecke, M. (2010). Alcohol as a risk factor for liver cirrhosis: A systematic review and meta-analysis. Drug and Alcohol Review, 29(4), 437–445. https://doi.org/10.1111/j.1465-3362.2009.00153.x

Reuter, M., & Fischl, B. (2011). Avoiding asymmetry-induced bias in longitudinal image processing. NeuroImage, 57(1), 19–21. https://doi.org/10.1016/j.neuroimage.2011.02.076

Reuter, M., Rosas, H. D., & Fischl, B. (2010). Highly accurate inverse consistent registration: A robust approach. NeuroImage, 53(4), 1181–1196. https://doi.org/10.1016/j.neuroimage.2010.07.020

Reuter, M., Schmansky, N. J., Rosas, H. D., & Fischl, B. (2012). Within-subject template estimation for unbiased longitudinal image analysis. NeuroImage, 61(4), 1402–1418. https://doi.org/10.1016/j.neuroimage.2012.02.084

Richard, G., Kolskår, K., Sanders, A.-M., Kaufmann, T., Petersen, A., Doan, N. T., Monereo Sánchez, J., Alnæs, D., Ulrichsen, K. M., Dørum, E. S., Andreassen, O. A., Nordvik, J. E., & Westlye, L. T. (2018). Assessing distinct patterns of cognitive aging using tissue-specific brain age prediction based on diffusion tensor imaging and brain morphometry. PeerJ, 6, e5908. https://doi.org/10.7717/peerj.5908

Richard, G., Kolskår, K., Ulrichsen, K. M., Kaufmann, T., Alnæs, D., Sanders, A.-M., Dørum, E. S., Monereo Sánchez, J., Petersen, A., Ihle-Hansen, H., Nordvik, J. E., & Westlye, L. T. (2020). Brain age prediction in stroke patients: Highly reliable but limited sensitivity to cognitive performance and response to cognitive training. NeuroImage: Clinical, 25, 102159. https://doi.org/10.1016/j.nicl.2019.102159

Ringen, P. A., Faerden, A., Antonsen, B., Falk, R. S., Mamen, A., Rognli, E. B., Solberg, D. K., Andreassen, O. A., & Martinsen, E. W. (2018). Cardiometabolic risk factors, physical activity and psychiatric status in patients in long-term psychiatric inpatient departments. Nordic Journal of Psychiatry, 72(4), 296–302. https://doi.org/10.1080/08039488.2018.1449012

Roalf, D. R., Quarmley, M., Elliott, M. A., Satterthwaite, T. D., Vandekar, S. N., Ruparel, K., Gennatas, E. D., Calkins, M. E., Moore, T. M., Hopson, R., Prabhakaran, K., Jackson, C. T., Verma, R., Hakonarson, H., Gur, R. C., & Gur, R. E. (2016). The impact of quality assurance assessment on diffusion tensor imaging outcomes in a large-scale population-based cohort. NeuroImage, 125, 903–919. https://doi.org/10.1016/j.neuroimage.2015.10.068

Roberson, L. L., Aneni, E. C., Maziak, W., Agatston, A., Feldman, T., Rouseff, M., Tran, T., Blaha, M. J., Santos, R. D., Sposito, A., Al-Mallah, M. H., Blankstein, R., Budoff, M. J., & Nasir, K. (2014). Beyond BMI: The “Metabolically healthy obese” phenotype & its association with clinical/subclinical cardiovascular disease and all-cause mortality -- a systematic review. BMC Public Health, 14(1), 14. https://doi.org/10.1186/1471-2458-14-14

Ronan, L., Alexander-Bloch, A. F., Wagstyl, K., Farooqi, S., Brayne, C., Tyler, L. K., & Fletcher, P. C. (2016). Obesity associated with increased brain age from midlife. Neurobiology of Aging, 47, 63–70. https://doi.org/10.1016/j.neurobiolaging.2016.07.010

Rosano, C., Marsland, A. L., & Gianaros, P. J. (2011). Maintaining Brain Health by Monitoring Inflammatory Processes: A Mechanism to Promote Successful Aging. Aging and Disease, 3(1), 16–33.

Rueckert, D., Sonoda, L. I., Hayes, C., Hill, D. L. G., Leach, M. O., & Hawkes, D. J. (1999). Nonrigid registration using free-form deformations: Application to breast MR images. IEEE Transactions on Medical Imaging, 18(8), 712–721. https://doi.org/10.1109/42.796284

Sanders, A.-M., Richard, G., Kolskår, K., Ulrichsen, K. M., Kaufmann, T., Alnæs, D., Beck, D., Dørum, E. S., Lange, A.-M. G. de, Nordvik, J. E., & Westlye, L. T. (2021). Linking objective measures of physical activity and capability with brain structure in healthy community dwelling older adults. MedRxiv, 2021.01.28.21250529. https://doi.org/10.1101/2021.01.28.21250529

Scott, K. M., McGee, M. A., Wells, J. E., & Oakley Browne, M. A. (2008). Obesity and mental disorders in the adult general population. Journal of Psychosomatic Research, 64(1), 97–105. https://doi.org/10.1016/j.jpsychores.2007.09.006

Sexton, C. E., Betts, J. F., Demnitz, N., Dawes, H., Ebmeier, K. P., & Johansen-Berg, H. (2016). A systematic review of MRI studies examining the relationship between physical fitness and activity and the white matter of the ageing brain. NeuroImage, 131, 81–90. https://doi.org/10.1016/j.neuroimage.2015.09.071

Smith, S. M. (2002). Fast robust automated brain extraction. Human Brain Mapping, 17(3), 143–155. https://doi.org/10.1002/hbm.10062

Smith, S. M., Jenkinson, M., Johansen-Berg, H., Rueckert, D., Nichols, T. E., Mackay, C. E., Watkins, K. E., Ciccarelli, O., Cader, M. Z., Matthews, P. M., & Behrens, T. E. J. (2006). Tract-based spatial statistics: Voxelwise analysis of multi-subject diffusion data. NeuroImage, 31(4), 1487–1505. https://doi.org/10.1016/j.neuroimage.2006.02.024

Smith, S. M., Jenkinson, M., Woolrich, M. W., Beckmann, C. F., Behrens, T. E. J., Johansen-Berg, H., Bannister, P. R., De Luca, M., Drobnjak, I., Flitney, D. E., Niazy, R. K., Saunders, J., Vickers, J., Zhang, Y., De Stefano, N., Brady, J. M., & Matthews, P. M. (2004). Advances in functional and structural MR image analysis and implementation as FSL. NeuroImage, 23, S208–S219. https://doi.org/10.1016/j.neuroimage.2004.07.051

Sone, D., Beheshti, I., Maikusa, N., Ota, M., Kimura, Y., Sato, N., Koepp, M., & Matsuda, H. (2019). Neuroimaging-based brain-age prediction in diverse forms of epilepsy: A signature of psychosis and beyond. Molecular Psychiatry. https://doi.org/10.1038/s41380-019-0446-9

Stanek, K. M., Grieve, S. M., Brickman, A. M., Korgaonkar, M. S., Paul, R. H., Cohen, R. A., & Gunstad, J. J. (2011). Obesity Is Associated With Reduced White Matter Integrity in Otherwise Healthy Adults*. Obesity, 19(3), 500–504. https://doi.org/10.1038/oby.2010.312

Strazzullo, P., D’Elia, L., Cairella, G., Garbagnati, F., Cappuccio, F. P., & Scalfi, L. (2010). Excess body weight and incidence of stroke: Meta-analysis of prospective studies with 2 million participants. Stroke, 41(5), e418–426. https://doi.org/10.1161/STROKEAHA.109.576967

Taki, Y., Kinomura, S., Sato, K., Inoue, K., Goto, R., Okada, K., Uchida, S., Kawashima, R., & Fukuda, H. (2008). Relationship Between Body Mass Index and Gray Matter Volume in 1,428 Healthy Individuals. Obesity, 16(1), 119–124. https://doi.org/10.1038/oby.2007.4

Taylor, J. R., Williams, N., Cusack, R., Auer, T., Shafto, M. A., Dixon, M., Tyler, L. K., Cam-CAN, & Henson, R. N. (2017). The Cambridge Centre for Ageing and Neuroscience (Cam-CAN) data repository: Structural and functional MRI, MEG, and cognitive data from a cross-sectional adult lifespan sample. NeuroImage, 144, 262–269. https://doi.org/10.1016/j.neuroimage.2015.09.018

Therkelsen, K. E., Pedley, A., Speliotes, E. K., Massaro, J. M., Murabito, J., Hoffmann, U., & Fox, C. S. (2013). Intramuscular fat and associations with metabolic risk factors in the Framingham Heart Study. Arteriosclerosis, Thrombosis, and Vascular Biology, 33(4), 863–870. https://doi.org/10.1161/ATVBAHA.112.301009

Tønnesen, S., Kaufmann, T., de Lange, A.-M. G., Richard, G., Doan, N. T., Alnæs, D., van der Meer, D., Rokicki, J., Moberget, T., Maximov, I. I., Agartz, I., Aminoff, S. R., Beck, D., Barch, D. M., Beresniewicz, J., Cervenka, S., Fatouros-Bergman, H., Craven, A. R., Flyckt, L., … Sellgren, C. (2020). Brain Age Prediction Reveals Aberrant Brain White Matter in Schizophrenia and Bipolar Disorder: A Multisample Diffusion Tensor Imaging Study. Biological Psychiatry: Cognitive Neuroscience and Neuroimaging, S2451902220301683. https://doi.org/10.1016/j.bpsc.2020.06.014

Tønnesen, S., Kaufmann, T., Doan, N. T., Alnæs, D., Córdova-Palomera, A., Meer, D. van der, Rokicki, J., Moberget, T., Gurholt, T. P., Haukvik, U. K., Ueland, T., Lagerberg, T. V., Agartz, I., Andreassen, O. A., & Westlye, L. T. (2018). White matter aberrations and age-related trajectories in patients with schizophrenia and bipolar disorder revealed by diffusion tensor imaging. Scientific Reports, 8(1), 14129. https://doi.org/10.1038/s41598-018-32355-9

van Buuren, S., & Groothuis-Oudshoorn, K. (n.d.). mice: Multivariate Imputation by Chained Equations in R. Journal of Statistical Software, 67.

Veraart, J., Fieremans, E., & Novikov, D. S. (2016). Diffusion MRI noise mapping using random matrix theory. Magnetic Resonance in Medicine, 76(5), 1582–1593. https://doi.org/10.1002/mrm.26059

Wagenmakers, E.-J., Lodewyckx, T., Kuriyal, H., & Grasman, R. (2010). Bayesian hypothesis testing for psychologists: A tutorial on the Savage–Dickey method. Cognitive Psychology, 60(3), 158–189. https://doi.org/10.1016/j.cogpsych.2009.12.001

Walther, K., Birdsill, A. C., Glisky, E. L., & Ryan, L. (2010). Structural brain differences and cognitive functioning related to body mass index in older females. Human Brain Mapping, 31(7), 1052–1064. https://doi.org/10.1002/hbm.20916

Ward, M. A., Carlsson, C. M., Trivedi, M. A., Sager, M. A., & Johnson, S. C. (2005). The effect of body mass index on global brain volume in middle-aged adults: A cross sectional study. BMC Neurology, 5(1), 23. https://doi.org/10.1186/1471-2377-5-23

Weinstein, G., Zelber-Sagi, S., Preis, S. R., Beiser, A. S., DeCarli, C., Speliotes, E. K., Satizabal, C. L., Vasan, R. S., & Seshadri, S. (2018). Association of Nonalcoholic Fatty Liver Disease With Lower Brain Volume in Healthy Middle-aged Adults in the Framingham Study. JAMA Neurology, 75(1), 97–104. https://doi.org/10.1001/jamaneurol.2017.3229

Willette, A. A., & Kapogiannis, D. (2015). Does the brain shrink as the waist expands? Ageing Research Reviews, 20, 86–97. https://doi.org/10.1016/j.arr.2014.03.007

Xu, J., Li, Y., Lin, H., Sinha, R., & Potenza, M. N. (2013). Body mass index correlates negatively with white matter integrity in the fornix and corpus callosum: A diffusion tensor imaging study. Human Brain Mapping, 34(5), 1044–1052. https://doi.org/10.1002/hbm.21491

Yusuf, S., Hawken, S., & Ounpuu, S. (2004). Effect of potentially modifiable risk factors associated with myocardial infarction in 52 countries (the INTERHEART study): Case-control study. ACC Current Journal Review, 13(12), 15–16. https://doi.org/10.1016/j.accreview.2004.11.072

