## Supplementary Material for "Adipose tissue distribution from body MRI is associated with cross-sectional and longitudinal brain age in adults"

**SI table 1.** Overview of Quality Assurance (QA) metrics for DTI data

| **QA metric abbreviation** | **Measure** |
|---|---|
| tsnr | Temporal-signal-to-noise-ratio |
| gmean | Global mean intensity |
| drift | Linear drift of signal over time |
| outmax | Outlier measurement maximum |
| outmean | Outlier measurement average |
| meanABSrmsb | Average absolute root-mean square |
| meanRELrmsb | Average relative root-mean square |
| maxABSrmsb | Maximum absolute root-mean square |
| maxRELrmsb | Maximum relative root-mean square |

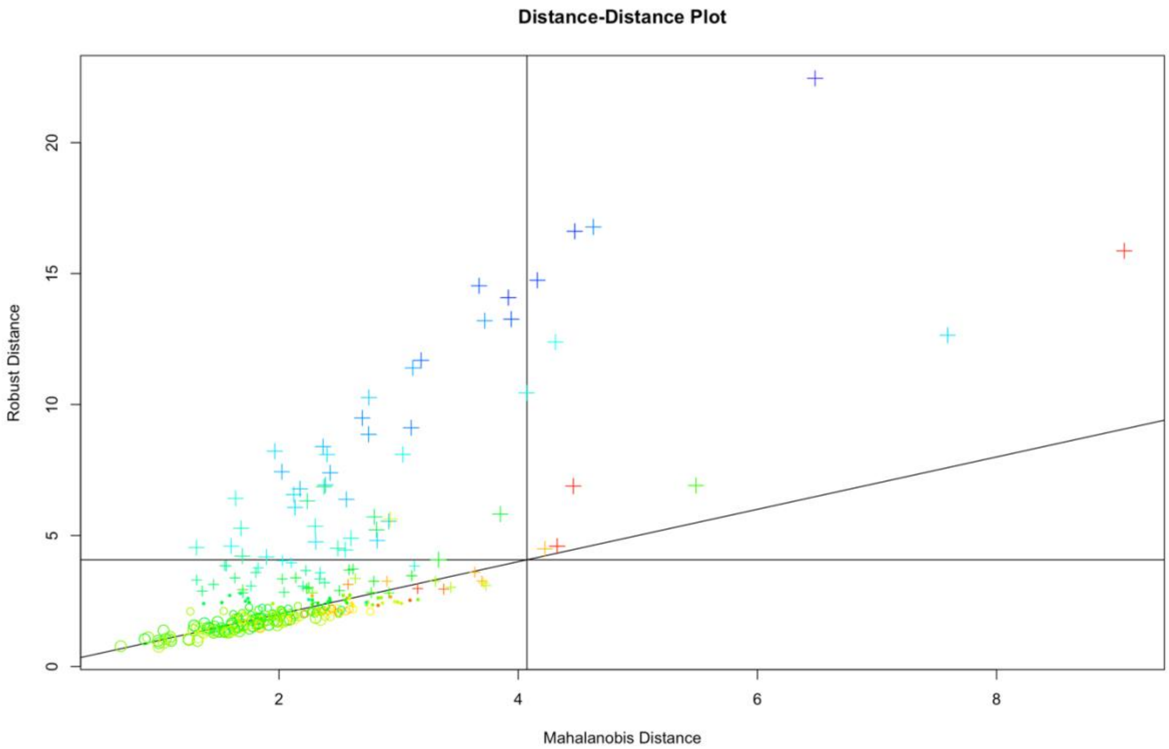


**SI Figure 1. Multivariate outlier detection algorithm.** Showing the Mahalanobis distance plot, which measures the distance between a point and a distribution to which that point belongs.

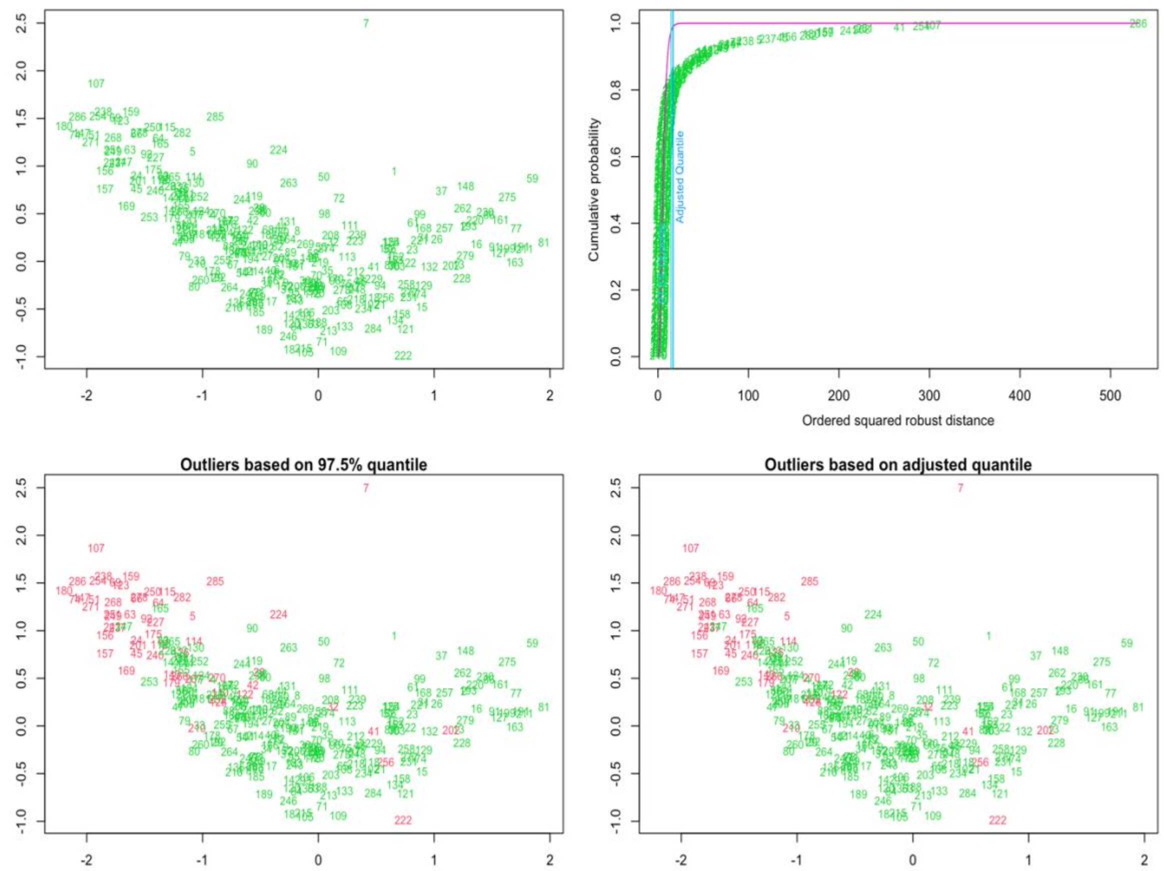


**SI Figure 2. Quantile and adjusted quantile.** Showing quantile plots which will solve for and order the squared Mahalanobis Distances for the given observations and plot them against the empirical Chi-Squared distribution function of these values. Every observation outside of the Chi-Square quantile is coloured in red.

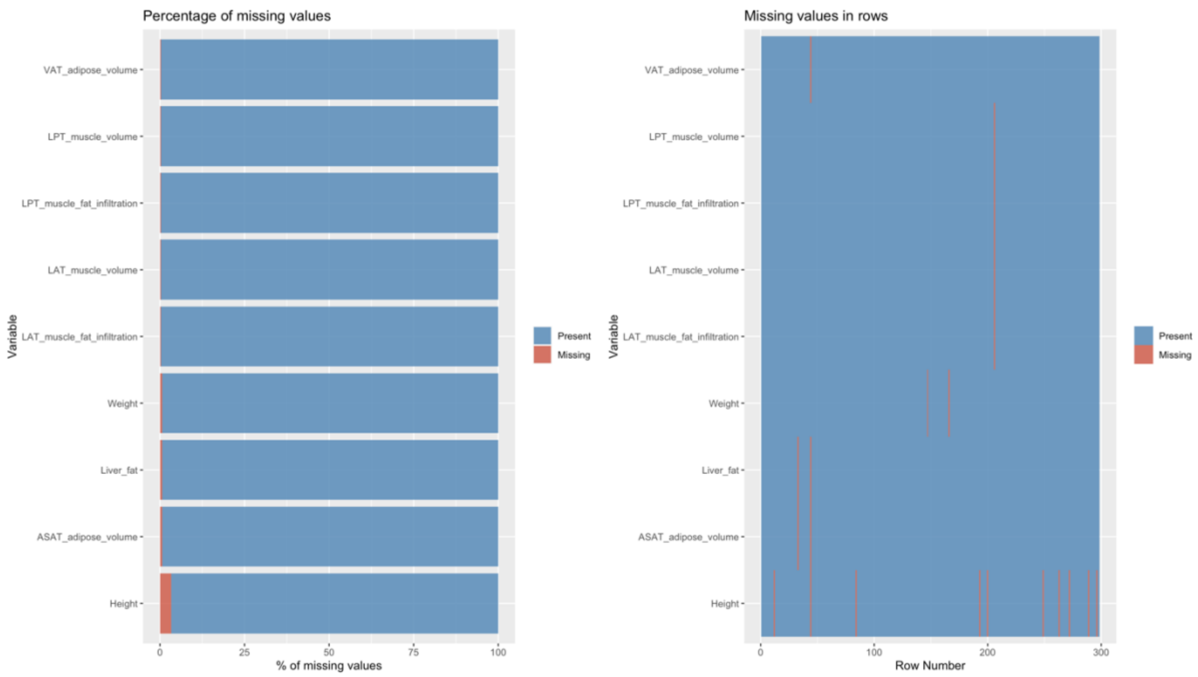

**SI Figure 3. Missing data report.** Showing percentage of missing data (left) and placement of missing data (right) for each adiposity measure prior to imputation.

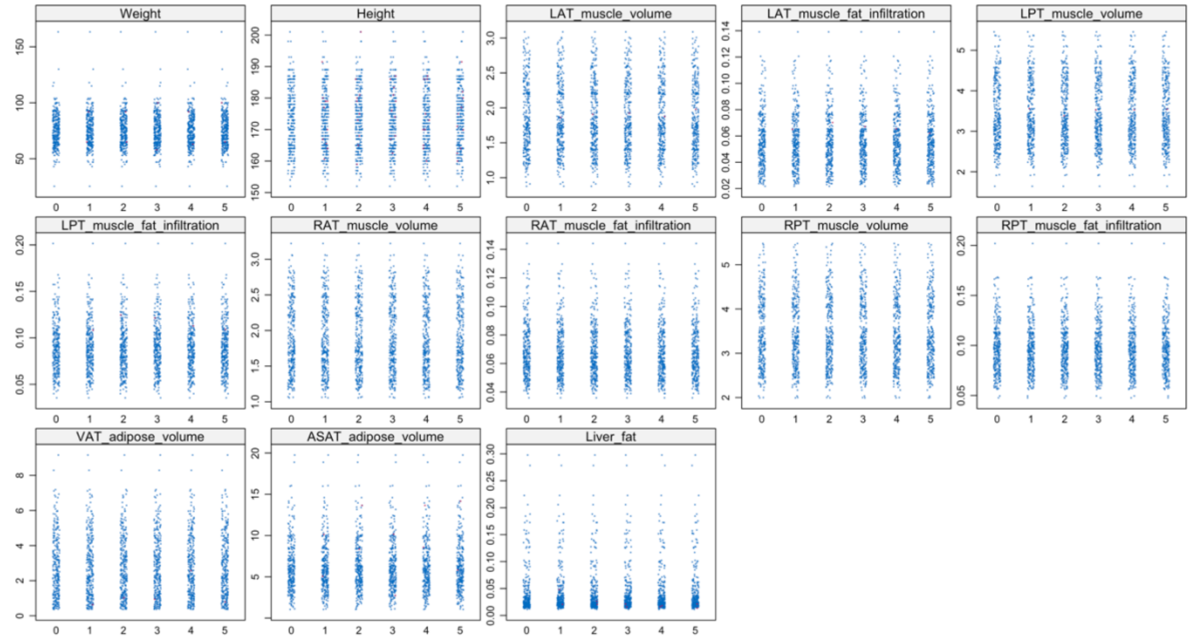


**SI Figure 4. MICE imputation.** Showing strip plot of imputed adiposity measures for five separate imputations. Blue dots represent original data, red dots represent imputed data.

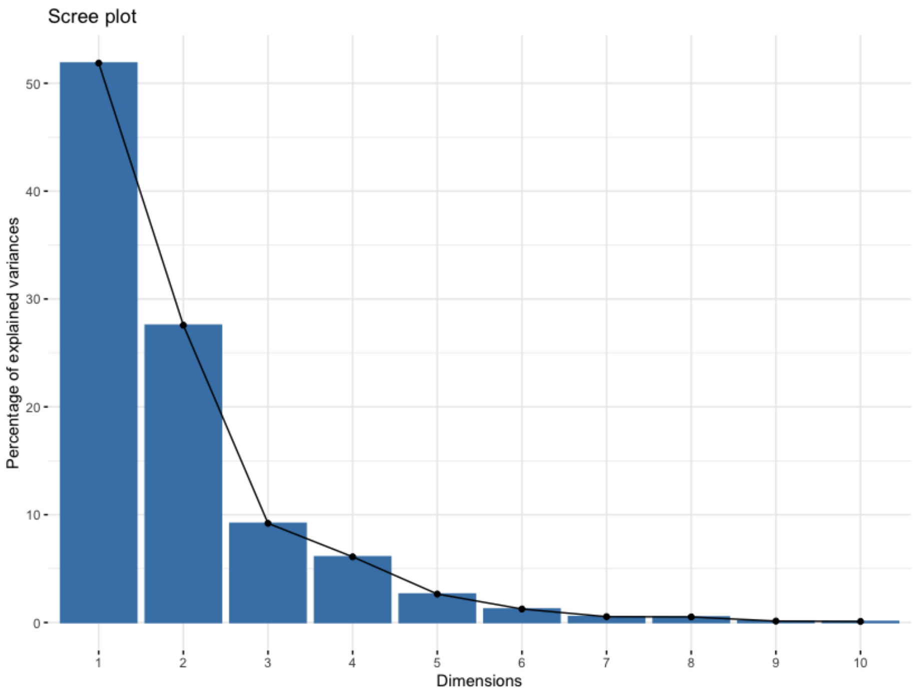


**SI Figure 5. Principle component analysis.** Scree plot showing percentage of explained variance for each dimension.

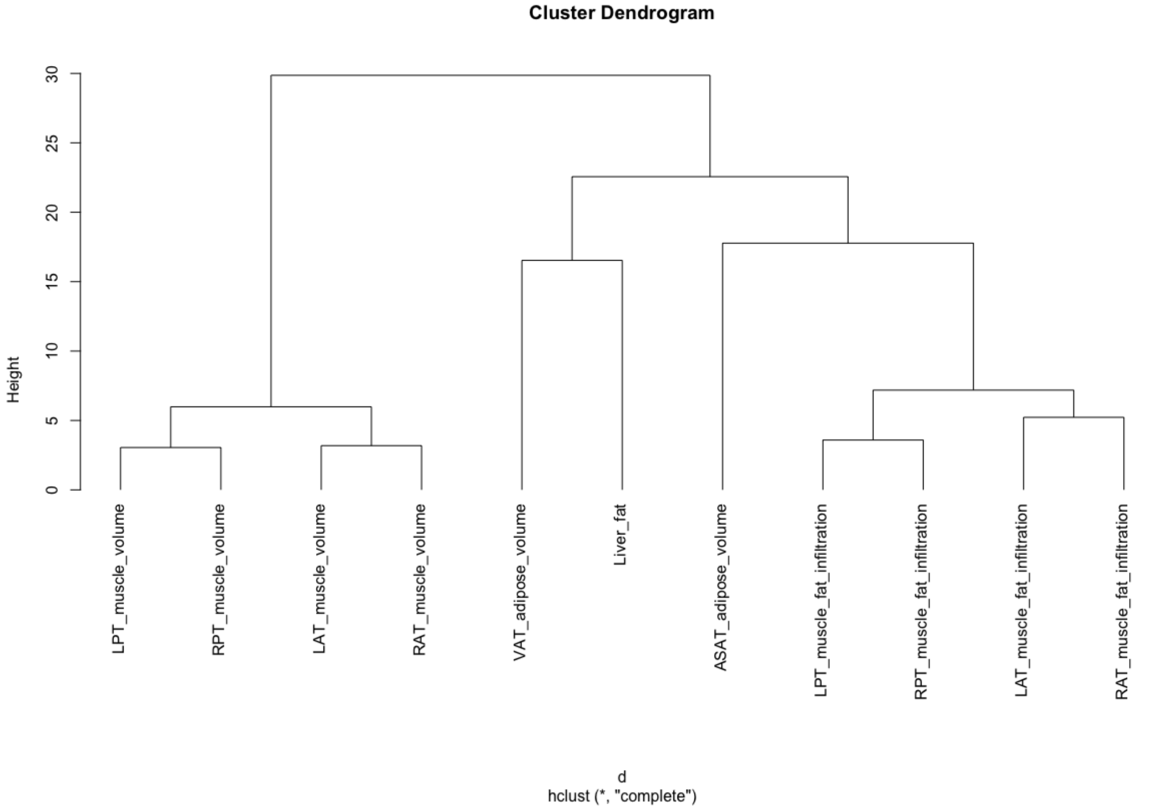


**SI Figure 6. Hierarchical clustering.** Showing results of 'hclust' using the complete linkage method to form hierarchical clusters.

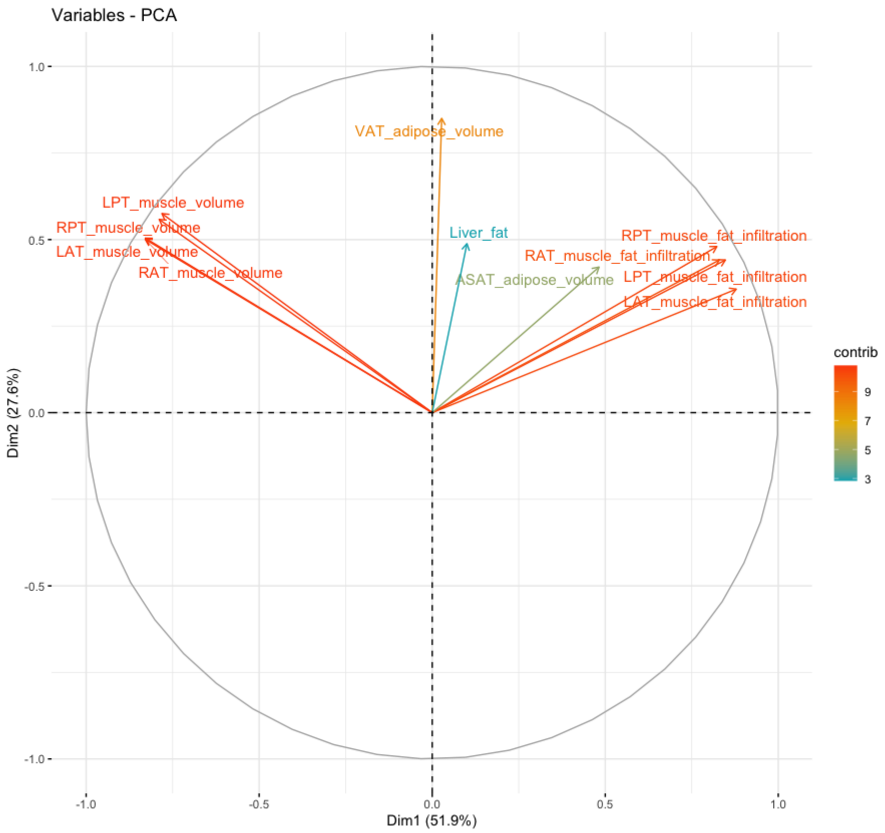

**SI Figure 7. Principle component correlations.** Showing directionality of correlations between adiposity variables, where positive correlated variables point to the same side and negative correlated variables point to opposites sides of the graph.

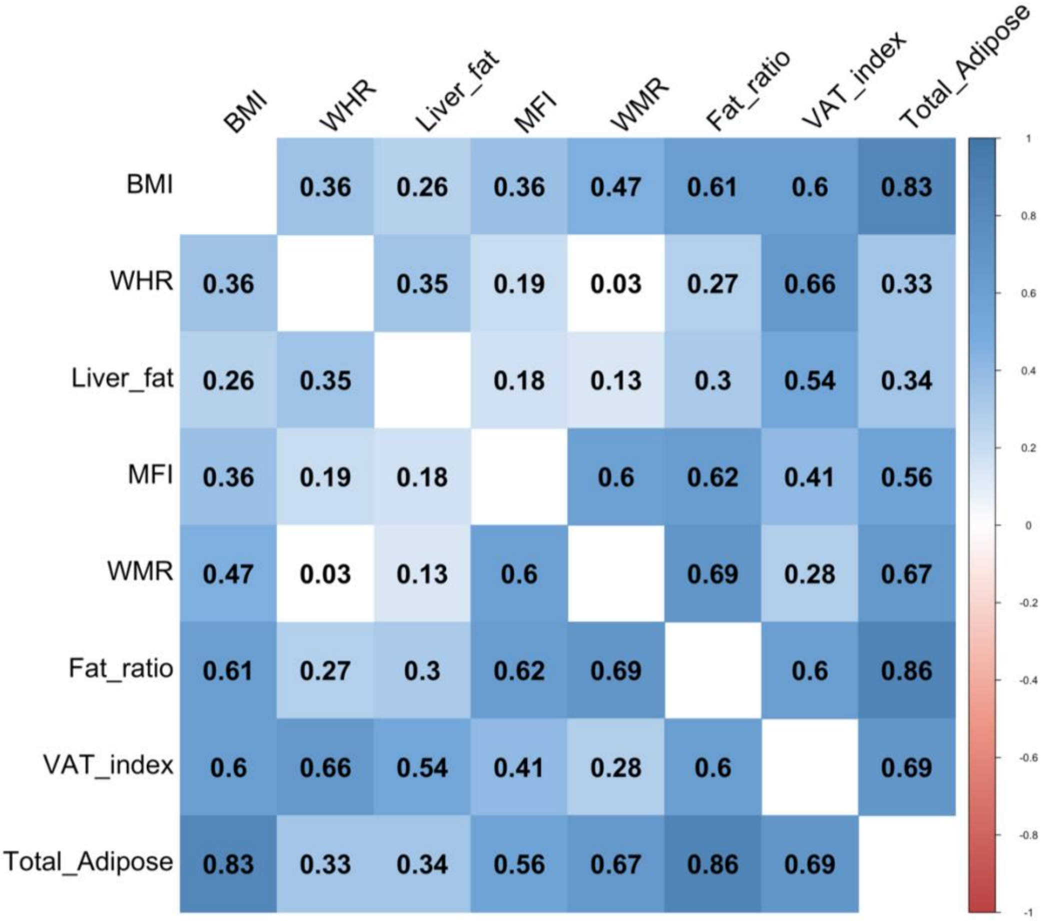


**SI Figure 8. Adiposity measure relatedness.** Correlation matrix showing relatedness of each adiposity variable.

**SI Table 2. Bayes Factor (BF).** Showing evidence ratio interpretations.

| Bayes factor $BF_{12}$ | | | Interpretation |
|---|---|---|---|
| | > | 100 | Extreme evidence for $M_1$ |
| 30 | - | 100 | Very Strong evidence for $M_1$ |
| 10 | - | 30 | Strong evidence for $M_1$ |
| 3 | - | 10 | Moderate evidence for $M_1$ |
| 1 | - | 3 | Anecdotal evidence for $M_1$ |
| | 1 | | No evidence |
| 1/3 | - | 1 | Anecdotal evidence for $M_2$ |
| 1/10 | - | 1/3 | Moderate evidence for $M_2$ |
| 1/30 | - | 1/10 | Strong evidence for $M_2$ |
| 1/100 | - | 1/30 | Very Strong evidence for $M_2$ |
| | < | 1/100 | Extreme evidence for $M_2$ |

**SI Table 3.** Average $R^2$, root mean square error (RMSE), and mean absolute error (MAE) ± standard deviation for the age prediction models within the training sample (Cam-CAN), test set, and age-corrected test set.

| | | Training sample (Cam-CAN) | Test set before age-bias correction | Test set after age-bias correction |
|---|---|---|---|---|
| DTI | $R^2$ | 0.82 ± 0.04 | 0.72 | 0.92 |
| | RMSE | 7.67 ± 0.83 | 10.11 | 5.12 |
| | MAE | 6.15 ± 0.55 | 8.37 | 4.06 |
| T1 | $R^2$ | 0.81 ± 0.04 | 0.73 | 0.87 |
| | RMSE | 7.93 ± 0.84 | 9.11 | 6.55 |
| | MAE | 6.19 ± 0.83 | 7.2 | 5.21 |

### Effects of age on adiposity measures

SI Figure 5 shows the posterior distributions for age on each adiposity measure. Full table of results for age effects on each variable can be seen in SI Table 4. Supplementary visualisation of the effects of age on a selection of adiposity measures can be seen in SI Figures 12.

Briefly, the tests revealed extreme evidence (BF < 0.01) in favour of an association between age and WMR ($\beta$ = 0.30), fat ratio ($\beta$ = 0.41), VAT index ($\beta$ = 0.43), WHR ($\beta$ = 0.47), and MFI ($\beta$ = 0.66). Very strong evidence was provided for total adiposity (BF = 0.06, $\beta$ = 0.25). The tests revealed strong evidence in favour of no association between age and BMI (BF = 10.87, $\beta$ = 0.05), while anecdotal evidence was provided for liver fat (BF = 1.58, $\beta$ = 0.16).

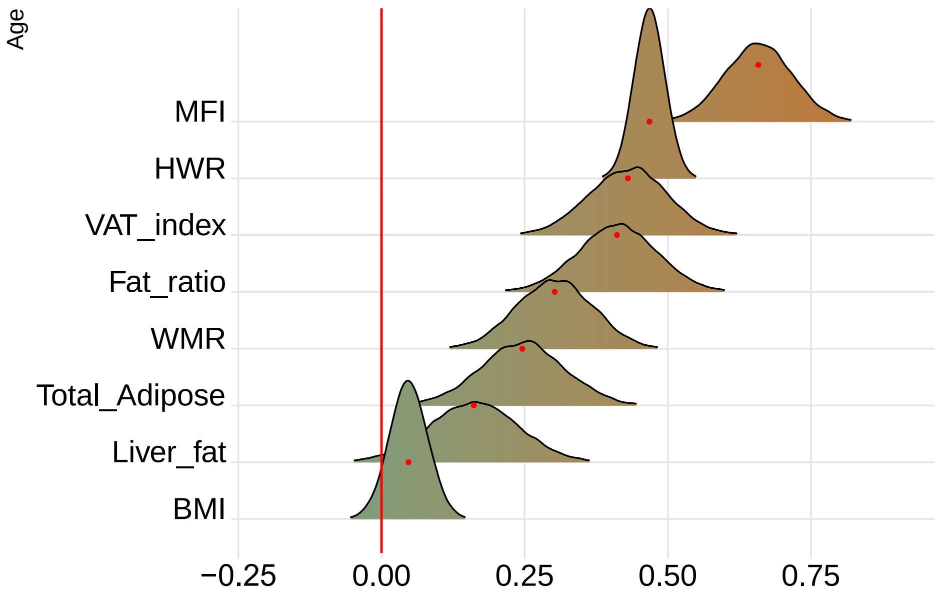


**SI Figure 9. Associations between adiposity and age.** The figure shows posterior distributions of the estimates of the coefficient. Estimates for age on each adiposity measure with red dot in each plot representing mean value. Colour scale follows direction of evidence. Width of distribution represents the uncertainty of the parameter estimates.

**SI Table 4. Associations between adiposity measures and age.** Variable ~ Age + Sex + Time (models = main effect).

| variable | predictor | estimate | lower95 | upper95 | p_higher_0 | p_lower_0 | evidence_0 |
|---|---|---|---|---|---|---|---|
| WMR | Age | 0.302 | 0.176 | 0.436 | 1 | 0 | 0 |
| Fat_ratio | Age | 0.411 | 0.277 | 0.549 | 1 | 0 | 0 |
| VAT_index | Age | 0.43 | 0.297 | 0.566 | 1 | 0 | 0 |
| Total_Adipose | Age | 0.246 | 0.105 | 0.397 | 1 | 0 | 0.06 |
| Liver_fat | Age | 0.161 | 0.012 | 0.315 | 0.98 | 0.02 | 1.58 |
| WHR | Age | 0.468 | 0.415 | 0.518 | 1 | 0 | 0 |
| BMI | Age | 0.047 | −0.017 | 0.113 | 0.925 | 0.075 | 10.87 |
| MFI | Age | 0.658 | 0.545 | 0.777 | 1 | 0 | 0 |

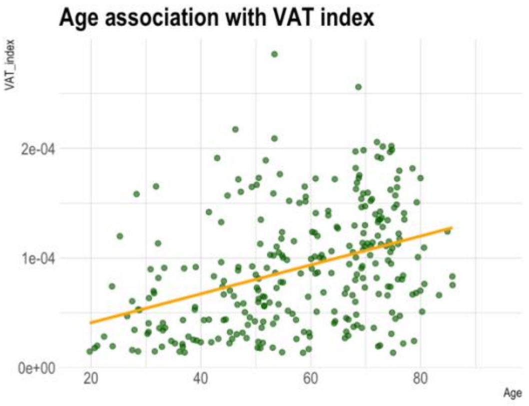


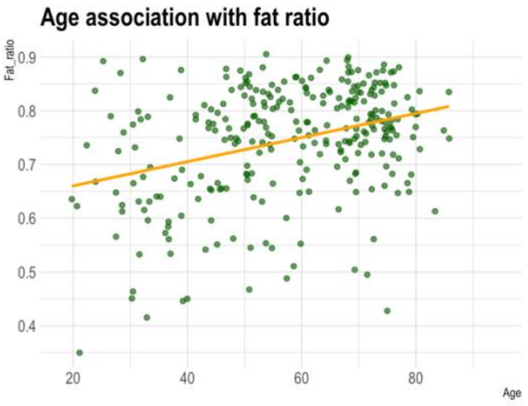

**SI Figure 10. Adiposity measures as functions of age.** Showing VAT Index and fat ratio.

**SI Table 5. Associations between BAGs and adiposity measures.** Showing main effect of each adiposity measure and interaction effects of adiposity measures and time and age.

| modality | cvr | estimate | lower95 | upper95 | p_higher_0 | p_lower_0 | evidence_0 | prior | model |
|---|---|---|---|---|---|---|---|---|---|
| BAG_DTI | WMR | –0.131 | –0.78 | 0.493 | 0.338 | 0.662 | 2.728 | 1 | Main effect |
| BAG_DTI | Fat_ratio | –0.495 | –1.116 | 0.116 | 0.057 | 0.943 | 0.914 | 1 | Main effect |
| BAG_DTI | Total_Adipose | –0.31 | –0.894 | 0.31 | 0.159 | 0.841 | 2.019 | 1 | Main effect |
| BAG_DTI | VAT_index | –0.048 | –0.658 | 0.557 | 0.437 | 0.563 | 3.196 | 1 | Main effect |
| BAG_DTI | Liver_fat | –0.133 | –0.719 | 0.46 | 0.329 | 0.671 | 2.902 | 1 | Main effect |
| BAG_DTI | WHR | 0.255 | –0.067 | 0.581 | 0.938 | 0.062 | 1.917 | 1 | Main effect |
| BAG_DTI | BMI | –0.211 | –0.519 | 0.081 | 0.084 | 0.916 | 2.619 | 1 | Main effect |
| BAG_DTI | MFI | 0.168 | –0.461 | 0.802 | 0.698 | 0.302 | 2.627 | 1 | Main effect |
| BAG_DTI | TP:WMR | 0.018 | –0.604 | 0.64 | 0.522 | 0.478 | 3.237 | 1 | TP-interaction |
| BAG_DTI | TP:Fat_ratio | –0.144 | –0.683 | 0.41 | 0.304 | 0.696 | 3.117 | 1 | TP-interaction |
| BAG_DTI | TP:Total_Adipose | –0.205 | –0.761 | 0.364 | 0.237 | 0.763 | 2.725 | 1 | TP-interaction |
| BAG_DTI | TP:VAT_index | –0.135 | –0.733 | 0.444 | 0.33 | 0.67 | 2.93 | 1 | TP-interaction |
| BAG_DTI | TP:Liver_fat | –0.213 | –0.914 | 0.434 | 0.267 | 0.733 | 2.513 | 1 | TP-interaction |
| BAG_DTI | TP:WHR | 0.389 | 0.068 | 0.71 | 0.991 | 0.009 | 0.444 | 1 | TP-interaction |
| BAG_DTI | TP:BMI | 0.174 | –0.124 | 0.48 | 0.869 | 0.131 | 3.434 | 1 | TP-interaction |
| BAG_DTI | TP:MFI | 0.43 | –0.279 | 1.135 | 0.88 | 0.12 | 1.406 | 1 | TP-interaction |

| | | | | | | | | | |
|---|---|---|---|---|---|---|---|---|---|
| BAG_DTI | Age:WMR | 0.301 | -0.36 | 0.955 | 0.814 | 0.186 | 1.921 | 1 | Age-interaction |
| BAG_DTI | Age:Fat_ratio | 0.176 | -0.44 | 0.769 | 0.716 | 0.284 | 2.75 | 1 | Age-interaction |
| BAG_DTI | Age:Total_Adipose | -0.014 | -0.6 | 0.585 | 0.481 | 0.52 | 3.197 | 1 | Age-interaction |
| BAG_DTI | Age:VAT_index | 0.223 | -0.455 | 0.895 | 0.741 | 0.259 | 2.299 | 1 | Age-interaction |
| BAG_DTI | Age:Liver_fat | 0.173 | -0.559 | 0.898 | 0.678 | 0.322 | 2.26 | 1 | Age-interaction |
| BAG_DTI | Age:WHR | 0.363 | 0.07 | 0.683 | 0.99 | 0.01 | 0.44 | 1 | Age-interaction |
| BAG_DTI | Age:BMI | 0.205 | -0.094 | 0.53 | 0.899 | 0.101 | 2.812 | 1 | Age-interaction |
| BAG_DTI | Age:MFI | 0.486 | -0.187 | 1.156 | 0.919 | 0.081 | 1.064 | 1 | Age-interaction |
| BAG_T1 | WMR | 0.775 | 0.001 | 1.551 | 0.976 | 0.024 | 0.402 | 1 | Main effect |
| BAG_T1 | Fat_ratio | -0.118 | -0.89 | 0.641 | 0.383 | 0.617 | 2.451 | 1 | Main effect |
| BAG_T1 | Total_Adipose | 0.088 | -0.675 | 0.824 | 0.593 | 0.407 | 2.501 | 1 | Main effect |
| BAG_T1 | VAT_index | 0.402 | -0.317 | 1.188 | 0.85 | 0.15 | 1.53 | 1 | Main effect |
| BAG_T1 | Liver_fat | 1.002 | 0.251 | 1.711 | 0.996 | 0.004 | 0.09 | 1 | Main effect |
| BAG_T1 | WHR | 0.173 | -0.232 | 0.602 | 0.794 | 0.206 | 3.4 | 1 | Main effect |
| BAG_T1 | BMI | -0.166 | -0.55 | 0.205 | 0.195 | 0.805 | 3.472 | 1 | Main effect |
| BAG_T1 | MFI | 0.917 | 0.118 | 1.663 | 0.989 | 0.011 | 0.168 | 1 | Main effect |
| BAG_T1 | TP:WMR | 0.675 | -0.076 | 1.402 | 0.962 | 0.038 | 0.569 | 1 | TP-interaction |
| BAG_T1 | TP:Fat_ratio | 0.341 | -0.34 | 1.029 | 0.832 | 0.168 | 1.717 | 1 | TP-interaction |
| BAG_T1 | TP:Total_Adipose | 0.341 | -0.359 | 1.041 | 0.832 | 0.168 | 1.791 | 1 | TP-interaction |
| BAG_T1 | TP:VAT_index | 0.408 | -0.31 | 1.146 | 0.865 | 0.135 | 1.493 | 1 | TP-interaction |

| | | | | | | | | | |
|---|---|---|---|---|---|---|---|---|---|
| BAG_T1 | TP:Liver_fat | 0.532 | -0.245 | 1.344 | 0.907 | 0.093 | 1.172 | 1 | TP-interaction |
| BAG_T1 | TP:WHR | 0.5 | 0.104 | 0.919 | 0.991 | 0.009 | 0.302 | 1 | TP-interaction |
| BAG_T1 | TP:BMI | 0.167 | -0.22 | 0.554 | 0.803 | 0.196 | 3.346 | 1 | TP-interaction |
| BAG_T1 | TP:MFI | 0.868 | 0.046 | 1.699 | 0.982 | 0.018 | 0.288 | 1 | TP-interaction |
| BAG_T1 | Age:WMR | 0.999 | 0.201 | 1.818 | 0.992 | 0.008 | 0.131 | 1 | Age-interaction |
| BAG_T1 | Age:Fat_ratio | 0.866 | 0.094 | 1.624 | 0.987 | 0.013 | 0.239 | 1 | Age-interaction |
| BAG_T1 | Age:Total_Adipose | 0.762 | 0.007 | 1.459 | 0.98 | 0.02 | 0.339 | 1 | Age-interaction |
| BAG_T1 | Age:VAT_index | 0.936 | 0.096 | 1.768 | 0.986 | 0.014 | 0.238 | 1 | Age-interaction |
| BAG_T1 | Age:Liver_fat | 0.937 | 0.093 | 1.859 | 0.98 | 0.02 | 0.247 | 1 | Age-interaction |
| BAG_T1 | Age:WHR | 0.683 | 0.29 | 1.076 | 1 | 0 | 0.01 | 1 | Age-interaction |
| BAG_T1 | Age:BMI | 0.42 | 0.025 | 0.822 | 0.98 | 0.02 | 0.618 | 1 | Age-interaction |
| BAG_T1 | Age:MFI | 1.034 | 0.201 | 1.828 | 0.993 | 0.007 | 0.119 | 1 | Age-interaction |

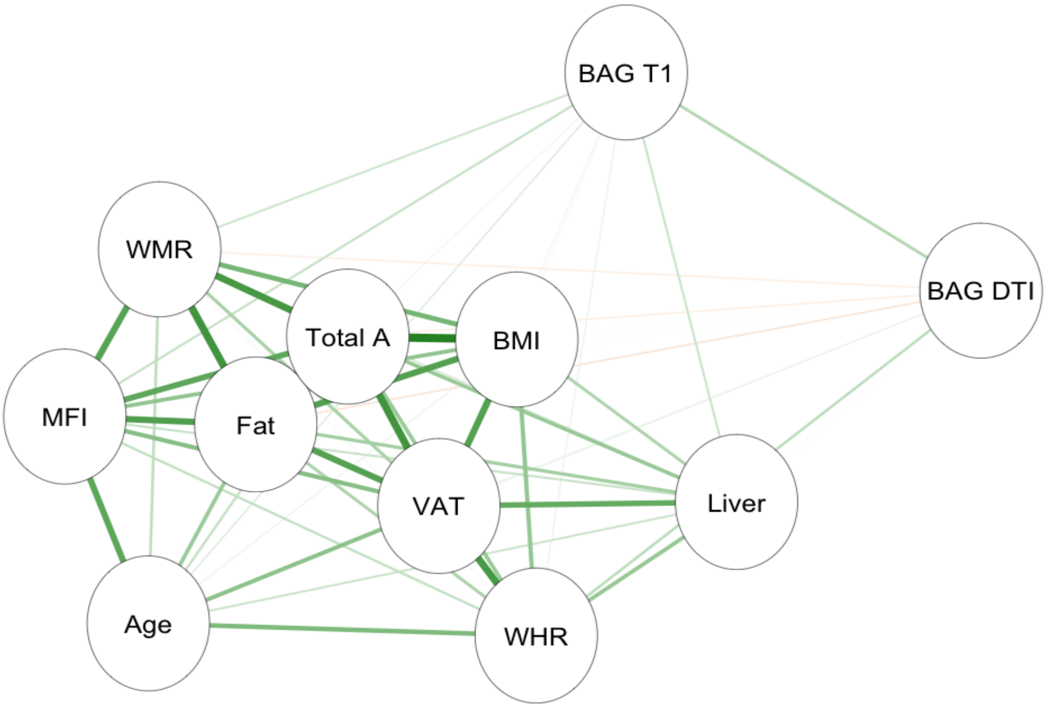


**SI Figure 11. Network correlation graph.** Showing correlations between adiposity measures and each BAG. The green lines indicate positive associations, and orange lines (none present) indicate negative associations. Strength of association marked by thickness of each line. Abbreviations: MFI – muscle fat infiltration; Fat – fat ratio; WHR – waist-to-hip ratio; VAT – visceral abdominal tissue index; WMR – weight-to-muscle ratio; Total A – total adipose; BMI – body-mass index; Liver – liver fat.

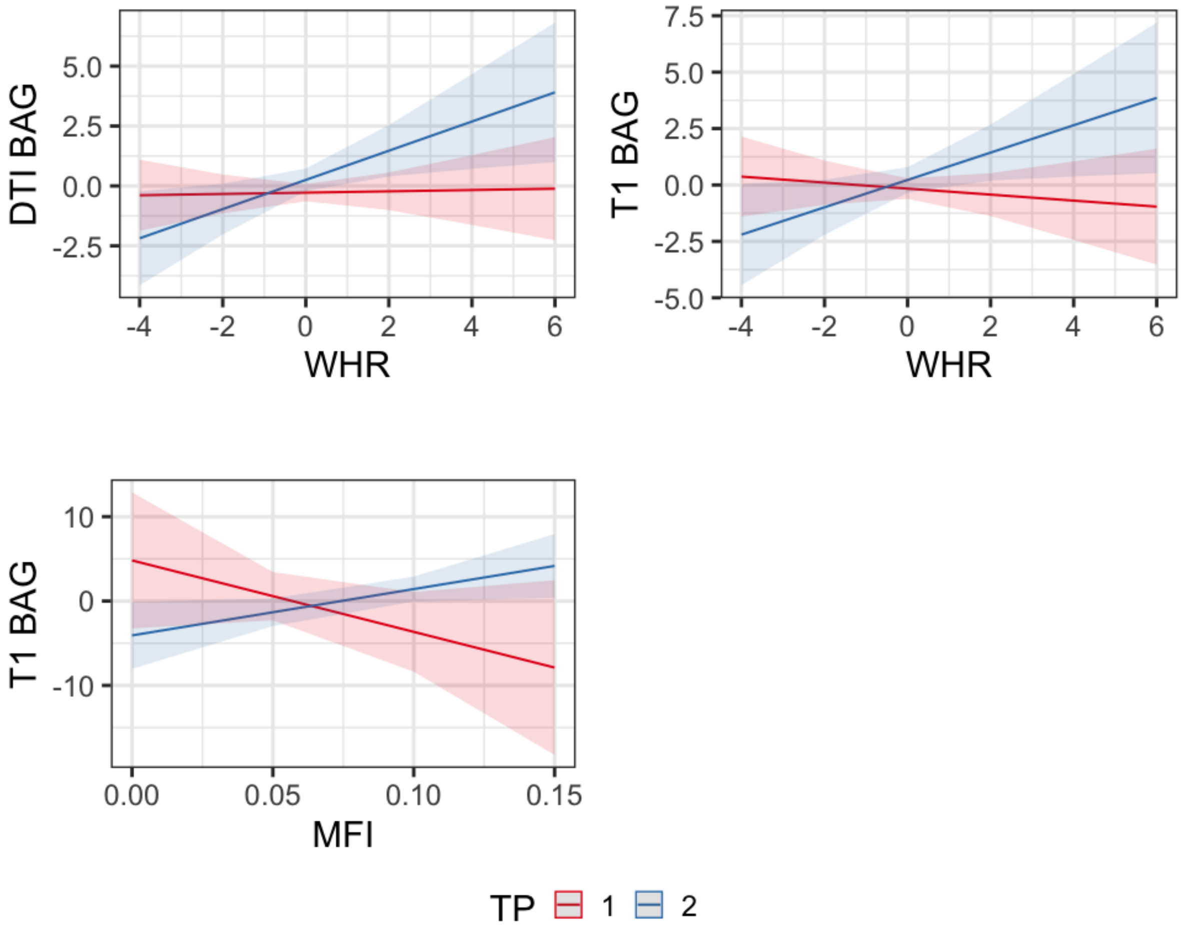


**SI Figure 12. Time interactions.** Interaction effects of waist-to-hip ratio and time on DTI BAG and T1 BAG, and muscle fat infiltration and time on T1 BAG. Models were fitted with BAGs as the dependent variable, adiposity measure as the independent variable (fixed effect), along with age, sex, with subject ID as random effects.
